# Experimental model of VZV reactivation reveals vaccine-induced mechanisms of protection against herpes zoster

**DOI:** 10.64898/2026.09.22.26363709

**Authors:** Michael J. Johnson, Thao Vu, Emily S. Ford, Lichen Jing, Kimberly Jordan, Mariah Brown, Shaobing Li, Troy Schedin, Megan Crotteau, Jennifer Canniff, Ashlynn Bennett, Neil Weaver, Kerry J. Laing, Seth Frietze, David M. Koelle, Myron J. Levin, Adriana Weinberg

**Author notes:** Address correspondence: Adriana Weinberg, MD, 12700 East 19th Ave., Room 11126, Aurora, CO 80045. Equally contributing first authors (Michael J. Johnson, Thao Vu, Emily S. Ford). Equally contributing senior authors (David M. Koelle, Myron J. Levin, Adriana Weinberg).

## Abstract

Herpes zoster (HZ), caused by varicella zoster virus (VZV) reactivation, is common in older adults with declining cell-mediated immune protection. A modestly efficacious live attenuated (ZVL) and a highly efficacious recombinant adjuvanted (RZV) zoster vaccine protect against HZ. To uncover the mechanism(s) associated with the superior efficacy of RZV, we developed a controlled human infection model consisting of intradermal administration of attenuated vOka VZV to older adults previously vaccinated with RZV or ZVL. Infiltration of VZV-specific CD4+ and CD8+ T cell clonotypes at the infection site was quicker and higher in RZV than ZVL recipients ≥5 years post-immunization due to the persistence of RZV-induced gE-reactive CD4+ and CD8+ T cell clonotypes in blood. High CD4+ cytotoxic T lymphocyte frequencies in blood on Day 1 post-inoculation predicted control of VZV replication. Persistence of circulating RZV-induced CD4+ and CD8+ CTL and rapid migration to sites of VZV replication are potential mechanisms underlying durable protection against HZ.

## Introduction

Herpes zoster (HZ) is a common painful and debilitating disease that affects one-third of unvaccinated people older than 60 years of age ^1^. Moreover, HZ has been associated with increased cardiovascular events, stroke, and dementia ^2,3^. HZ is caused by the varicella zoster virus (VZV), which causes a self-limited primary infection during childhood (aka varicella or chickenpox). During this infection, immune competent children develop a robust immune response that controls VZV replication and promotes resolution of the disease ^4^. However, during varicella, VZV establishes latency in sensory ganglia and subsequently periodically reactivates. Most reactivations are asymptomatic, being controlled by cell-mediated immune (CMI) responses ^5–15^. However, when VZV-specific CMI decreases to some critical level because of aging or immune compromise, the reactivated VZV propagates sufficiently to cause HZ ^1^.

Although the exact mechanism of protection against HZ is incompletely understood, two vaccines were developed to boost VZV-CMI and prevent HZ and/or its complications. The first vaccine, zoster vaccine live (ZVL), consisted of the attenuated vOka strain of VZV. ZVL had limited efficacy of 70% in the 1^st^ year after immunization in 50-60-year-old adults, but the efficacy declined significantly as the age of the vaccinee increased ^16,17^. Moreover, the efficacy of ZVL waned rapidly within 5 years and disappeared by 10 years ^18^. In contrast, the currently recommended recombinant zoster vaccine (RZV), which contains the recombinant VZV glycoprotein E (VZV-gE) and the AS01B adjuvant, has an efficacy higher than 90% in the first 3 years after immunization that is minimally affected by the age of the vaccinee and persists at 72% 10 years after immunization ^19^. We sought to explain the prominent clinical differences between the two vaccines by determining the immunologic mechanisms underlying their divergent efficacy and identifying immune correlates of protection against VZV replication. To achieve this goal, we developed an experimental model of controlled cutaneous VZV infection designed to mimic VZV reactivation.

Controlled human infection models (CHIM) for viral, bacterial and parasitic pathogens, including influenza, respiratory syncytial virus, and *S. pneumoniae*, have been used to identify the immune correlates of protection against these infections ^20^. The biology of VZV, which does not infect laboratory animals, was previously studied in skin organoid cultures and in humanized mouse models with human skin allografts ^4^ but these models are not suitable for the characterization of VZV protective immune responses against HZ. Previous investigators used VZV antigen intradermal injections to study local immune responses to VZV in seropositive individuals ^21^ but, in the absence of viral replication, an immune correlate of protection could not be determined. Our CHIM of VZV reactivation is based on knowledge that reactivation of latent VZV DNA in sensory ganglia results in VZV traveling antegrade through neuronal axons to the dermal-epidermal junction in the skin ^1^. We mimic this stage of reactivation by inoculating the participants intradermally with the vOka VZV strain in ZVL. This procedure generates a limited VZV skin infection at the dermal/intradermal junction, which is the site of VZV replication of the translocated reactivated VZV in HZ. This model is based on previous studies demonstrating that Intradermal inoculation of ZVL is safe and immunogenic ^22^, resulting in immune responses remarkably similar to those occurring during HZ ^23,24^.

The current study showed that both RZV and ZVL recipients developed robust systemic and local immune responses shortly after the viral challenge. We also demonstrated distinctive immune responses between recipients of the two vaccines, and between recent and remote vaccine recipients, which may explain the different efficacies of ZVL and RZV and provide additional insight into their mechanisms of immune protection against HZ.

## Results

### Characteristics of the study population

The study enrolled 105 participants without prior HZ or immune deficiency divided into the following 4 vaccine groups: received ZVL ≥5 years before entry (<u>Remote ZVL</u>; N=18); received RZV ≥5 years before entry (<u>Remote RZV</u>, N=20); received ZVL 6±1 months before entry (<u>Recent ZVL</u>; N=32); and concluded RZV immunization 6±1 months before entry (<u>Recent</u> <u>RZV</u>; N=35). At entry, participants were median 61 years of age, including 66 women and 92 White non-Hispanic individuals (**Figure 1B**). Participants in the <u>Remote</u> vaccine groups were older than those in the <u>Recent</u> vaccine groups, but other demographic characteristics were balanced. All participants received a VZV vOka challenge at entry in the form of intradermal ZVL and had blood collected pre-challenge (Day 0) and at Days 1, 3, and 7 post-challenge (**Figure 1A**). In addition, at the timepoints mentioned above, subsets of 3-5 participants per group in the <u>Remote ZVL</u> and <u>Remote RZV</u> groups had skin biopsies obtained from the site of VZV intradermal inoculation (**Figure 1A**).

**Figure 1.**
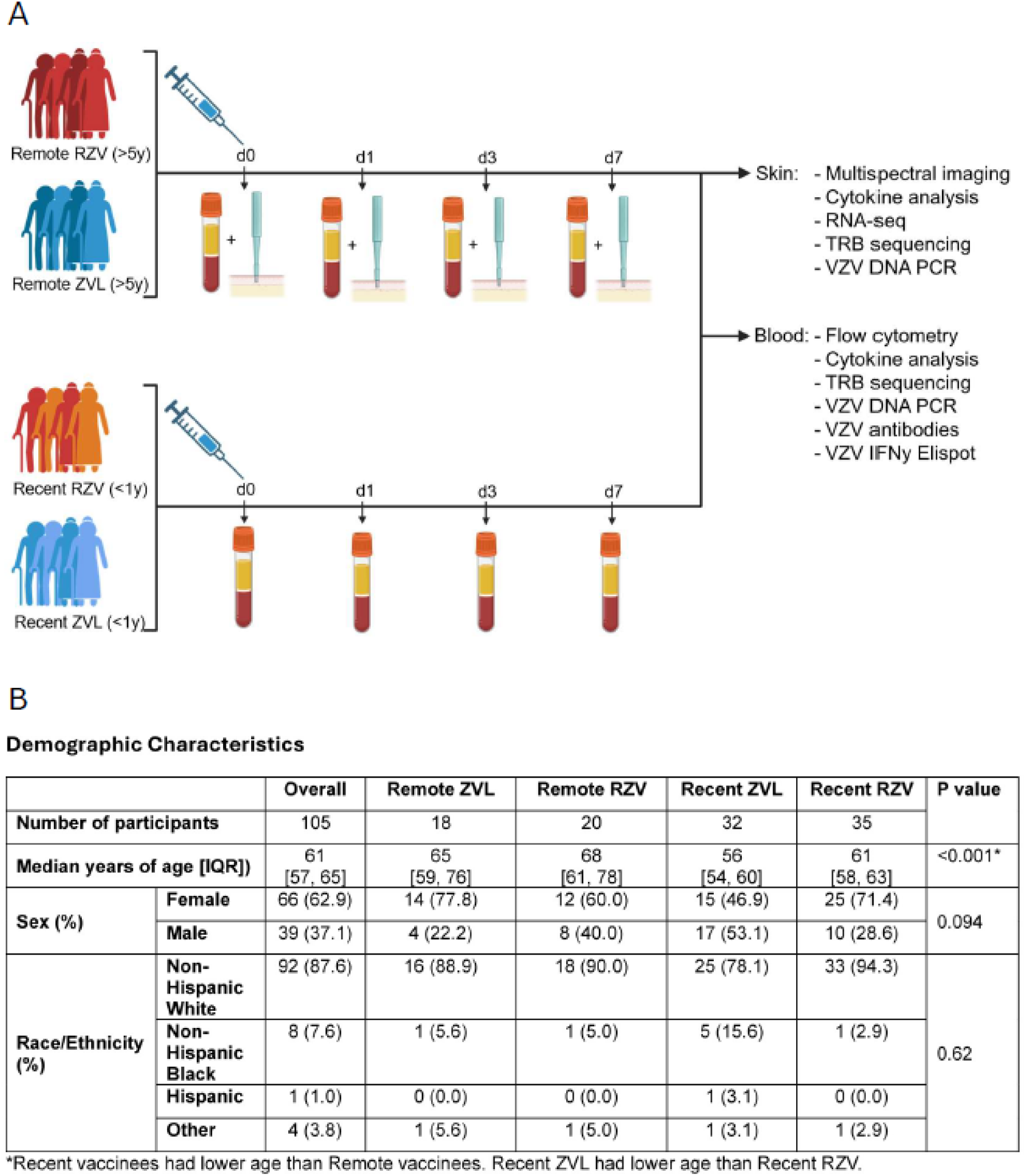
Study design and demographic characteristics of the study population. Panel. **A** shows a schematic representation of the study design. Participants were divided into 4 groups as indicated on the graph. All participants received an intradermal challenge with vOka symbolized by the syringe and had samples collected and assays performed as shown on the graph. **Panel B** shows the demographic characteristics of the participants overall and in each group.

### VZV vOka replication and recall immune responses to the viral challenge

VZV DNA was detected by PCR in all skin fragments obtained post-challenge. In addition, 27 out of a total of 36 skin biopsies had suitable RNA for sequencing and analysis of VZV transcripts. VZV mRNA were absent in 8 Day 0 biopsies but were present in 5 out of 5 Day 1 biopsies tapering to 1 out of 6 fragments on Day 7 (**Figure 2A**). The transcripts included both early and late genes, such as ORF 14 (gC), 17, 31 (gB), 37 (gH), 60 (gL), 67 (gE), etc., indicating that the injected VZV underwent complete cycles of replication. There were no appreciable differences between the <u>Remote ZVL</u> and <u>Remote RZV</u> groups in VZV transcription in this small sample of biopsies.

**Figure 2.**
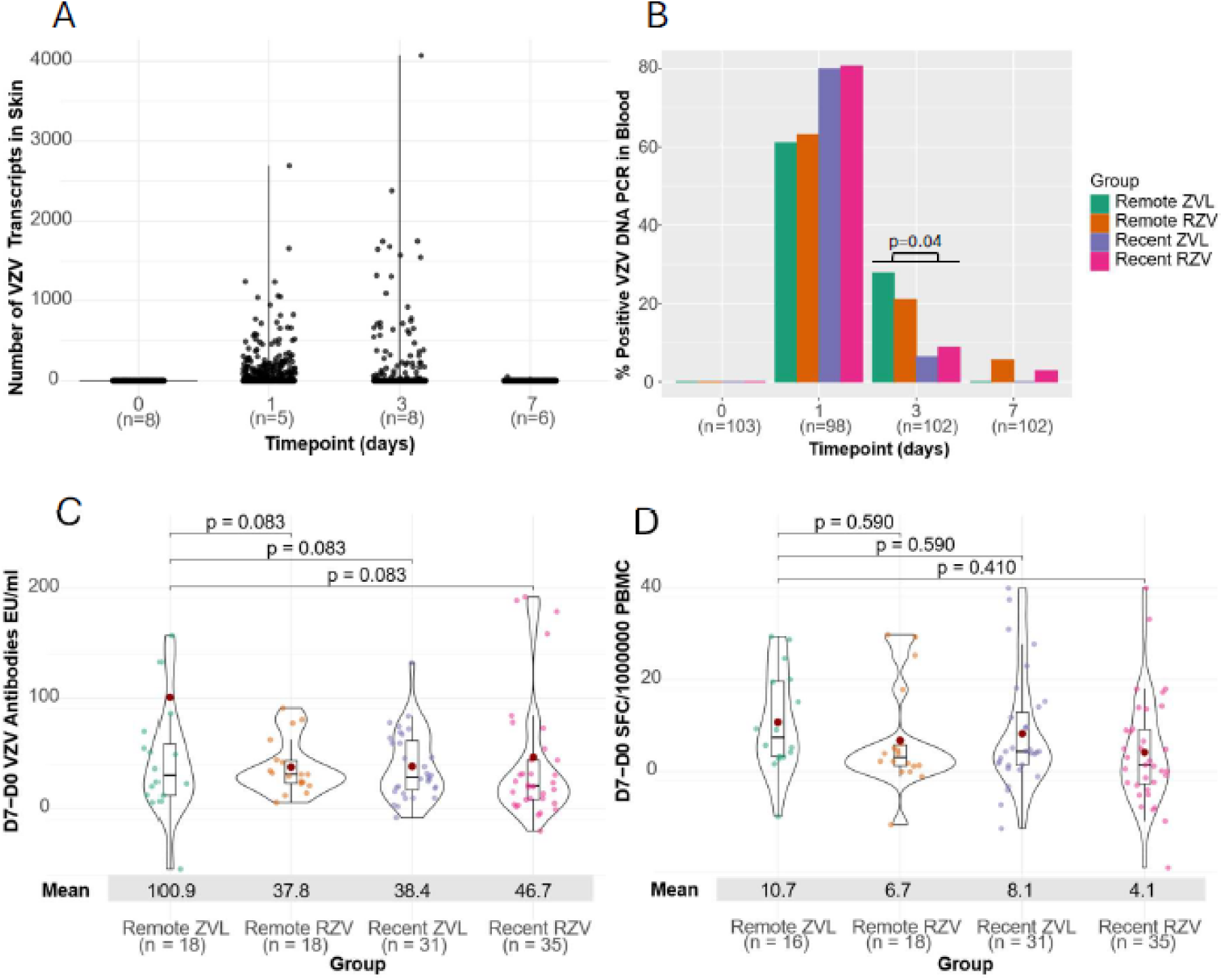
VZV vOka replication and adaptive immune responses. Panel. **A** shows the number of VZV mRNA copies in biopsy homogenates at each timepoint before and after inoculation of VZV vOka attenuated strain. Data were derived from 27 biopsies with suitable RNA for this analysis obtained from Remote ZVL and Remote RZV recipients. Each dot represents a transcript. Transcripts include VZV immediate early, early and structural genes. **Panel B** shows the proportions of participants in each group with detectable VZV DNAemia before and after VZV vOka challenge. Data were derived from a total of 105 participants with N of participants contributing information at each timepoint specified on the graph. p value shows the comparison between Remote and Recent vaccinees using chi-square test. **Panel C** shows the VZV antibody increase from pre-challenge to 7 days post-challenge in each study group. Data were derived from 102 participants with paired blood samples. The dots in the center of the violin plots indicate means. The rectangle and whiskers show medians, upper and lower quartiles and range. p values are FDR-adjusted and show pairwise comparisons using ANOVA. **Panel D** shows the increase from pre-challenge to 7 days post-challenge in IFNγ+ VZV spot-forming cells/10^6^ PBMC measured by ELISPOT in each study group. Data were derived from 102 participants with paired blood samples. p values are FDR-adjusted and show pairwise comparisons using ANOVA.

We postulated that replicating virus entering the blood stream could be detected as VZV DNA by PCR (paired samples of plasma and white blood cells) as a surrogate marker of viremia. VZV is exceedingly difficult to isolate from blood and DNAemia has been accepted as a surrogate^14^. Blood was obtained on Days 0, 1, 3, and 7 from all participants who came for the scheduled visits. VZV DNA was detected in 0 of 103 samples tested pre-inoculation; 71 of 98 samples (72.5%) on Day 1; 14 of 102 samples (14%) on Day 3; and 2 of 102 samples (2%) on Day 7 (**Figure 2B**). The comparison across the 4 vaccine groups did not reveal significant differences in the proportions of positive results at any visit (p ≥ 0.11). However, participants in the <u>Remote ZVL and Remote RZV</u> groups combined had higher frequency of VZV DNAemia on Day 3 than participants in the <u>Recent ZVL</u> and <u>Recent RZV</u> groups combined (24% vs. 8%; p=0.04), consistent with waning of protective immunity over time in <u>Remote vaccine</u> recipients.

To further characterize VZV vOka replication we measured the adaptive immune responses to the challenge, which we previously showed to increase with the severity of HZ ^23^ and, presumably, with the amount of virus produced, as demonstrated in other infections ^25^. Anti-VZV antibodies were measured on Days 0 and Day 7 post-challenge. Participants in all vaccine groups showed robust increases in antibody concentrations from pre- to post-VZV challenge (**Figure 2C**). The comparison across groups revealed more than 2-fold higher antibody increments from pre- to post-challenge in the <u>Remote ZVL</u> group compared with the other three groups. Although the difference reached only marginal statistical significance (FDR-adjusted p=0.08), this finding suggested an immune reaction to higher VZV replication in the <u>Remote ZVL</u> group. VZV-CMI measured by IFNγ ELISPOT increased in all groups from Day 0 to Day 7 without significant differences across groups (**Figure 2D**).

### Kinetics of the early systemic immune responses to VZV intradermal inoculation

We measured plasma levels of the chemokines MCP-1, MIP-1α, MIP-1β; cytokines produced mainly by myeloid cells (IFN-α2a, IL-6, IL-12p70, IL-15); cytokines produced mainly by T cells (GM-CSF, IL-2, IL-17A); and cytokines produced both by innate and adaptive immune cells (IFN-γ, IL-10, TNF-α) using a microarray chemiluminescence assay on plasma collected on Days 0, 1, 3, and 7. We used flow cytometry to define the immune cell response detectable in blood. CCR7 and CD45RO were used to characterize CD4+ and CD8+ T cell differentiation, including naïve (T_NAIVE_), stem cell memory (T_SCM_), central memory (T_CM_), effector memory (T_EM_), and terminally differentiated effectors (T_EMRA_). We measured in vivo activation (CD38+HLA-DR+), proliferation (Ki 67+), cytotoxicity (CD107a+) and IFN-γ production of T cells and NK cells. In vivo activation of total, classical, intermediate and nonclassical monocytes, total DC, cDC1, cDC2, phagocytic or inflammatory (CD16+) cDC, and pDC were assessed by expression of CD40, TNF-α, PD-L1, and IL-10. Gating strategy is shown in **Supplementary Figure 1**.

To determine the overall kinetics of the immune response to viral replication, we performed a pooled analysis that combined the data from all participants at each timepoint regardless of vaccine group. The pooled analysis of the cytokines and chemokines on Days 0, 1, 3, and 7 showed significantly higher levels (FDR-adjusted p<0.05 by ANOVA for repeated measures) of IFN-γ, IL-6, IL-10, MCP-1 and TNF-α on Day 1 post-challenge compared to Day 0 and of IL-15 on Days 1 and 3 post-challenge (**Figure 3A; Supplementary Figure 2**).

Monocytes and all DC subsets showed significantly increased CD40 and/or PD-L1 expression on Days 1 and/or 3 (**Figure 3B; Supplementary Figure 2**). NK expression of CD107a, CD38 and HLA-DR, IFN-γ, and Ki 67 peaked on Day 3 post-challenge (**Figure 3C, Supplementary Figure 2**). CD4+ T cell activation, proliferation and T_EMRA_ differentiation increased up to Day 7, while CD8+ T_EMRA_ differentiation peaked earlier, on Day 3 (**Figures 3D and 3E, Supplementary Figure 2**).

**Figure 3.**
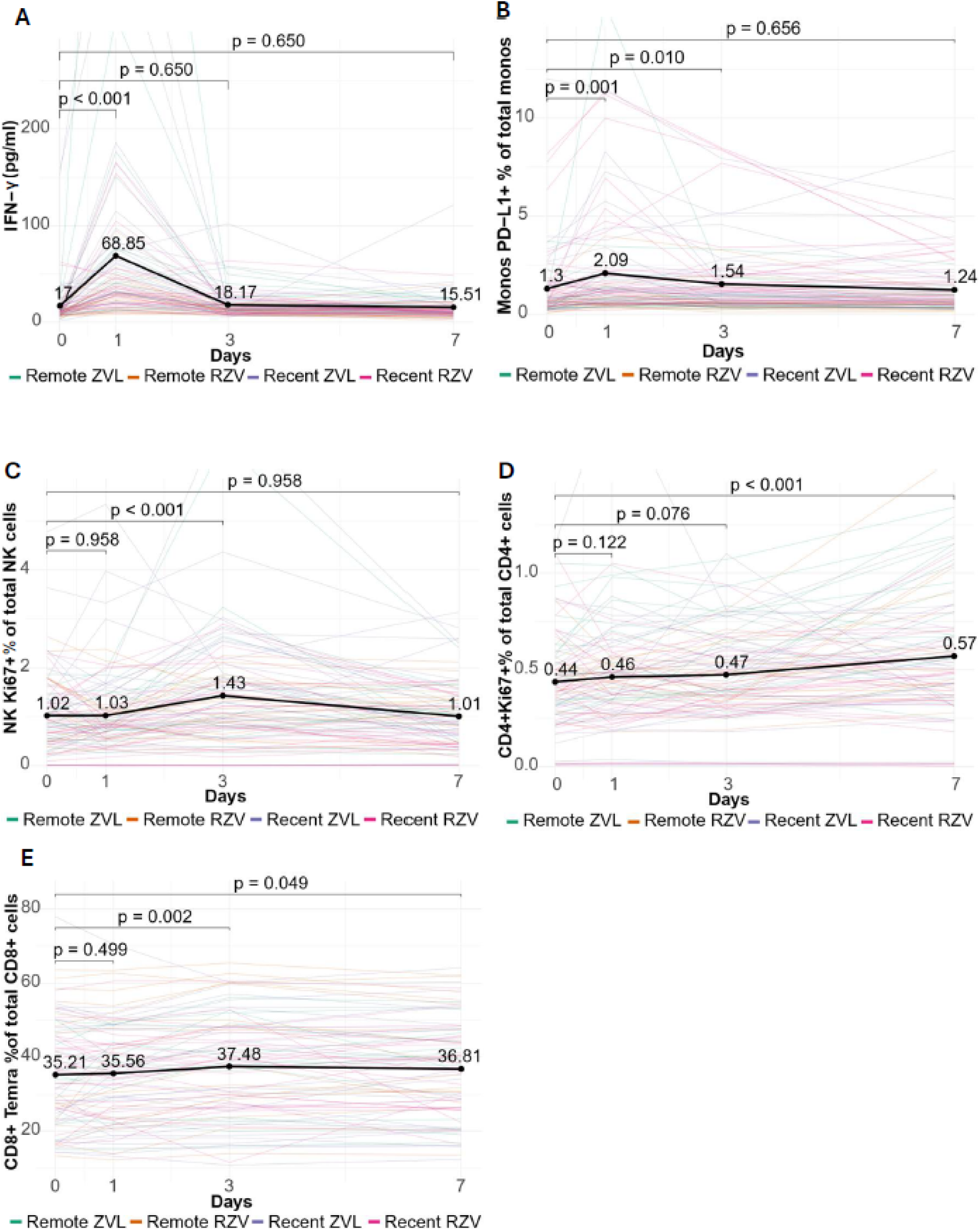
Kinetics of the systemic immune responses to VZV replication. Data were derived from 88 participants with blood samples at each timepoint. Fine lines represent each participant and are individually color-coded by study group. Some scales do not include the highest values, as indicated by the truncated fine lines, to optimize visualization of the differences across timepoints. The thick black line shows the average across all participants. The dots and numbers on the thick black line represent means. The bars show FDR-adjusted p values for pair-wise comparisons from Day 0 to subsequent timepoints. **Panels A, B, C, D** and **E** show typical representations of cytokine, APC, NK cell, CD4+ and CD8+ T cell significant responses, respectively, identified by ANOVA for repeated values with FDR p<0.05. Remaining significant responses are shown in **Supplementary** Figure 2.

The difference in responses to the viral challenge among vaccine groups was studied by comparing the increase in cytokine/chemokine levels and activated or differentiated immune cells from pre-challenge (Day 0) to Days 1, 3, and 7 post-challenge for each group. Cytokine and chemokine plasma levels showed significantly higher increases from Day 0 to Day 3 and/or Day 7 in the <u>Remote ZVL</u> group compared to other groups, indicating persistence of the inflammatory response up to Day 7 in this group. Differences included TNF-α on Day 3; MCP-1 on Days 3 and 7; and IL-6 on Day 7 (**Figure 4A, Supplementary Figure 3**). Myeloid cell activation did not significantly differ across groups. The comparison of the NK cells showed a significantly higher increase of in vivo IFN-γ expression from Day 0 to Day 1 in the <u>Recent RZV</u> group compared to the other groups (**Figure 4B**). We observed multiple significant differences in the CD4+ and CD8+ T cell in vivo responses across groups. (**1**) <u>Recent RZV</u> recipients displayed the highest increases in CD8+CD107a+ cytotoxic T lymphocytes (CTL) on Day 1 and of CD4+CD107a+ CTL and CD8+IFN-γ+ Th1 on Day 3. Notably, T cell differences reached statistical significance with the <u>Remote</u> and <u>Recent ZVL</u> groups, but not with the <u>Remote RZV</u> group (**Figure 4D, Supplementary Figure 3**), suggesting that the differences might have been driven by the type of vaccine. (**2**) <u>Remote RZV</u> recipients showed the highest increase in CD8+CD38+HLADR+ T_NAIVE_ on Day 7 (**Supplementary Figure 3**). (**4**) <u>Remote ZVL</u> participants had the highest CD4+CD38+HLADR+ T_EMRA_ elevations on Day 1 and highest CD4+Ki67+ proliferating cells on Day 7 (**Figure 4C, Supplementary Figure 3**). Collectively, the comparisons among groups revealed that <u>Recent RZV</u> recipients had the most rapid response and that <u>Remote ZVL</u> recipients had the most prolonged inflammation, possibly reflecting extended viral replication.

**Figure 4.**
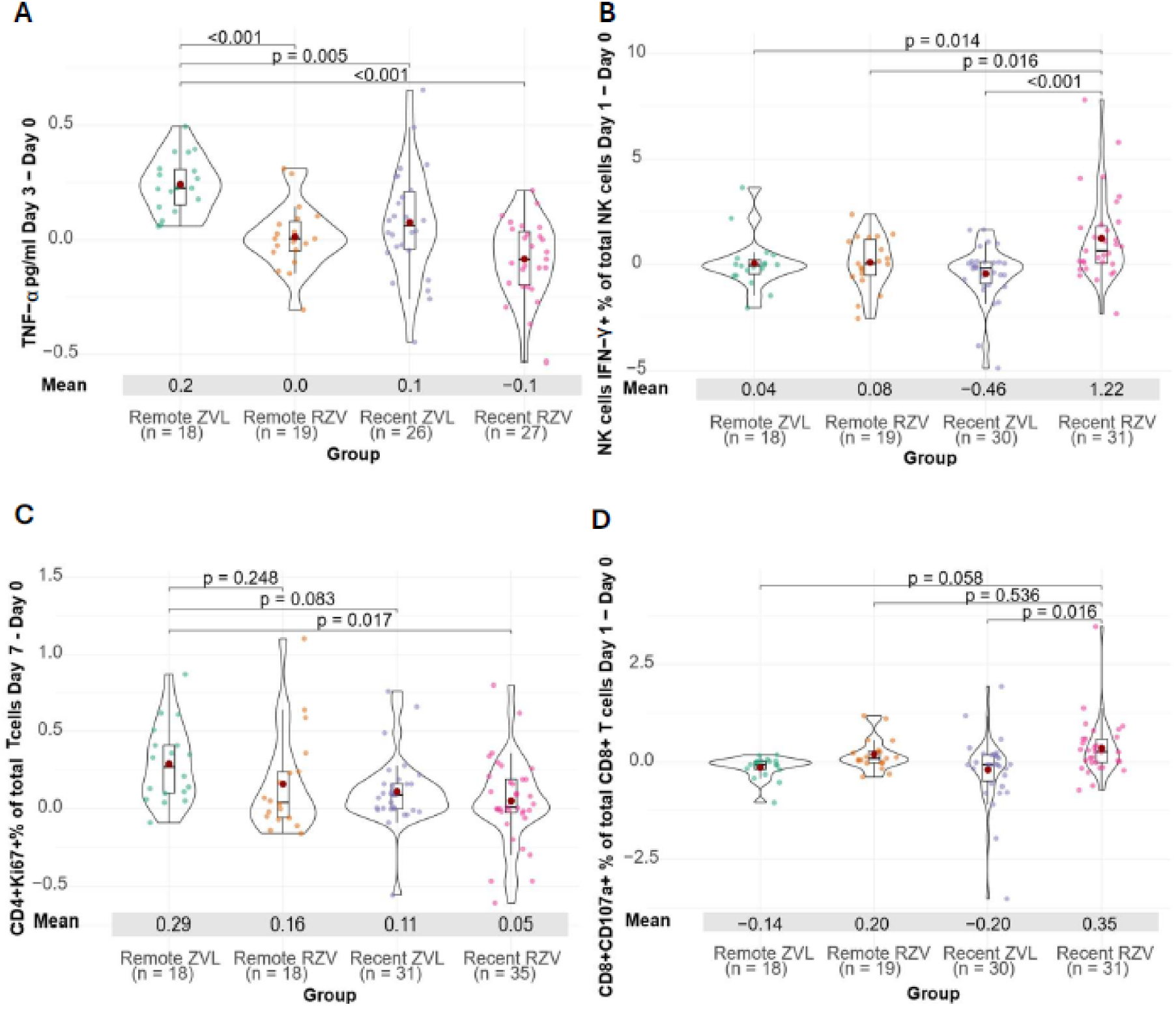
Differential systemic immune responses to VZV vOka replication among vaccine groups. Increases in blood immune parameters from pre-challenge to Days 1, 3 and 7 post-challenge were compared among vaccine groups using data from 102 participants with paired samples. The timepoints for which differences are shown are indicated on the y axis of each graph. The dots represent individual differences from pre-challenge to the timepoint indicated on the y axis in each participant. In the center of the violin plots, the boxes show medians, quartiles and ranges and the brown dots indicate means. The numeric value of the means is shown under the x axis. p values shown on the graphs were calculated by post-hoc pairwise comparisons between groups. Analytes with significant differences across groups were identified using ANOVA with FDR-adjusted p < 0.05. The group with the highest responses is used as the reference for pairwise comparisons. **Panels A, B, C,** and **D** show typical representations of cytokine, NK cell, CD4+ and CD8+ T cell differences, respectively, across groups. Overall, the <u>Remote ZVL</u> group showed higher increases at late timepoints compared with other groups, while the <u>Recent RZV</u> group showed higher increases at early timepoints. Other comparisons with significant differences across groups are shown in **Supplementary** Figure 3.

### Systemic immunologic predictors of control of VZV replication post-challenge

We used VZV DNAemia on Day 3 post-challenge, which was detected in 14 of 102 participants, as a surrogate measure of the magnitude of VZV replication. Out of all the cytokine, chemokine, and cell subsets measured in blood pre- and post-challenge, including pre-challenge anti-VZV antibody concentrations and IFN-γ ELISPOT, only three parameters were significantly associated with the outcome measure (**Figure 5A**). The frequency of CD4+CD107a+ CTL in blood on Day 1 had a significant negative association with Day 3 VZV DNAemia (effect estimate= -24.3, FDR-adjusted p=0.04), suggesting that CD4+ CTL play a role in the control of viral replication. The frequency of VZV DNAemia on Day 3 showed positive associations with MCP-1 concentrations in plasma and with the frequency of circulating CD8+ T_EM_ on Day 3, but with low effect sizes (effect estimate of 0.1 and 0.02, respectively, FDR-adjusted p of 0.049 and 0.045, respectively). The positive association suggested that VZV replication drove these immune responses.

**Figure 5.**
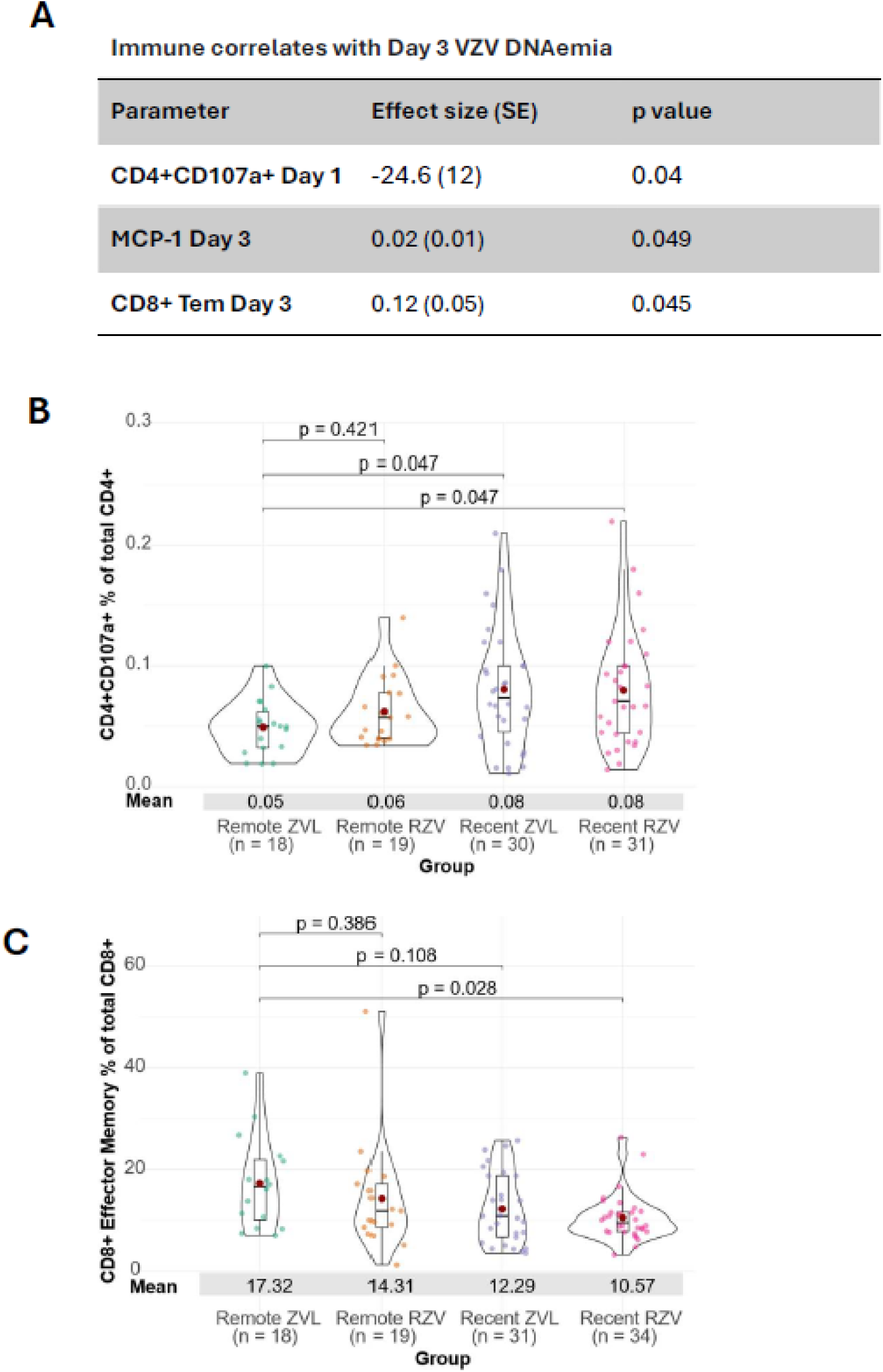
Immune correlates with viral replication. **Panel A** shows the immune parameters significantly correlated with the incidence of VZV DNAemia on Day 3, their effect sizes on the outcome measure and p values. **Panels B** and **C** show the distribution across groups of the parameters significantly associated with viral replication, which also showed significant differences across the vaccine groups. The strength of the differences across groups is shown by the adjusted p values calculated by ANOVA. The immune parameter analyzed is shown on the y axis and the number of participants contributing data to each analysis is shown on the x axis.

Next, we determined if the vaccine groups differed with respect to the magnitude of the parameters associated with viral replication. The comparison across groups of CD4+CD107a+ CTL on Day 1 post-challenge showed higher frequencies in <u>Recent RZV</u> and <u>Recent ZVL</u> recipients compared to <u>Remote ZVL</u> recipients (**Figure 5B**). In contrast, the frequency of CD8+ T_EM_ on Day 3 was higher in the <u>Remote ZVL</u> than in the <u>Recent RZV</u> group (**Figure 5C**). Collectively, these observations are consistent with the waning of ZVL-conferred immune protection over time, and relatively similar protection conferred by the two vaccines in the first 6 months after immunization.

### Local immune responses to VZV infection

The skin inflammatory responses post-VZV inoculation were assessed on the 36 punch biopsies described in Figure 1A. The specimens were analyzed using bulk RNA-seq (same specimens analyzed for VZV mRNA transcription shown above), and cytokine and chemokine measurements in tissue homogenates, and immunofluorescence in formalin-fixed sections.

#### Skin transcription profiles elicited by the VZV vOka intradermal challenge

were delineated in samples with suitable RNA, which consisted of 15 biopsies in the <u>Remote ZVL</u> group and 12 biopsies in the <u>Remote RZV</u> group. Significant changes (FDR-adjusted p<0.05) in the local transcription profiles started on Day 1 post-inoculation in both groups with immune activation and progressed to deactivating the immune response and tissue remodeling on Day 7 (**Figure 6A**). Vaccine-specific changes included interferon-driven antiviral responses on Day 1 in the <u>Remote ZVL</u> group with upregulation of IRF7, GSDMA, VAV3, DROSHA, and BPIFA2, followed by regulatory signaling with increased TGFβ transcription and decreased OAS2 and BACH2 on Day 3 and Day 7. In contrast, the <u>Remote RZV</u> group showed chemokine-driven immune recruitment with upregulation of CCL23 on Days 1 and 3, and increased lymphocyte immune signaling with upregulation of RAG1, mir181C, ZKSCAN3 and PLCB1 on Day 1; followed by immune regulation with NLRP2 downregulation and NKAPL upregulation on Day 3, and NKAPL, mir182, and mir200C upregulation on Day 7. Both groups showed increased expression of SEMA3A and RGS19 on Days 3 and 7 consistent with immune down-regulation and tissue remodeling.

**Figure 6.**
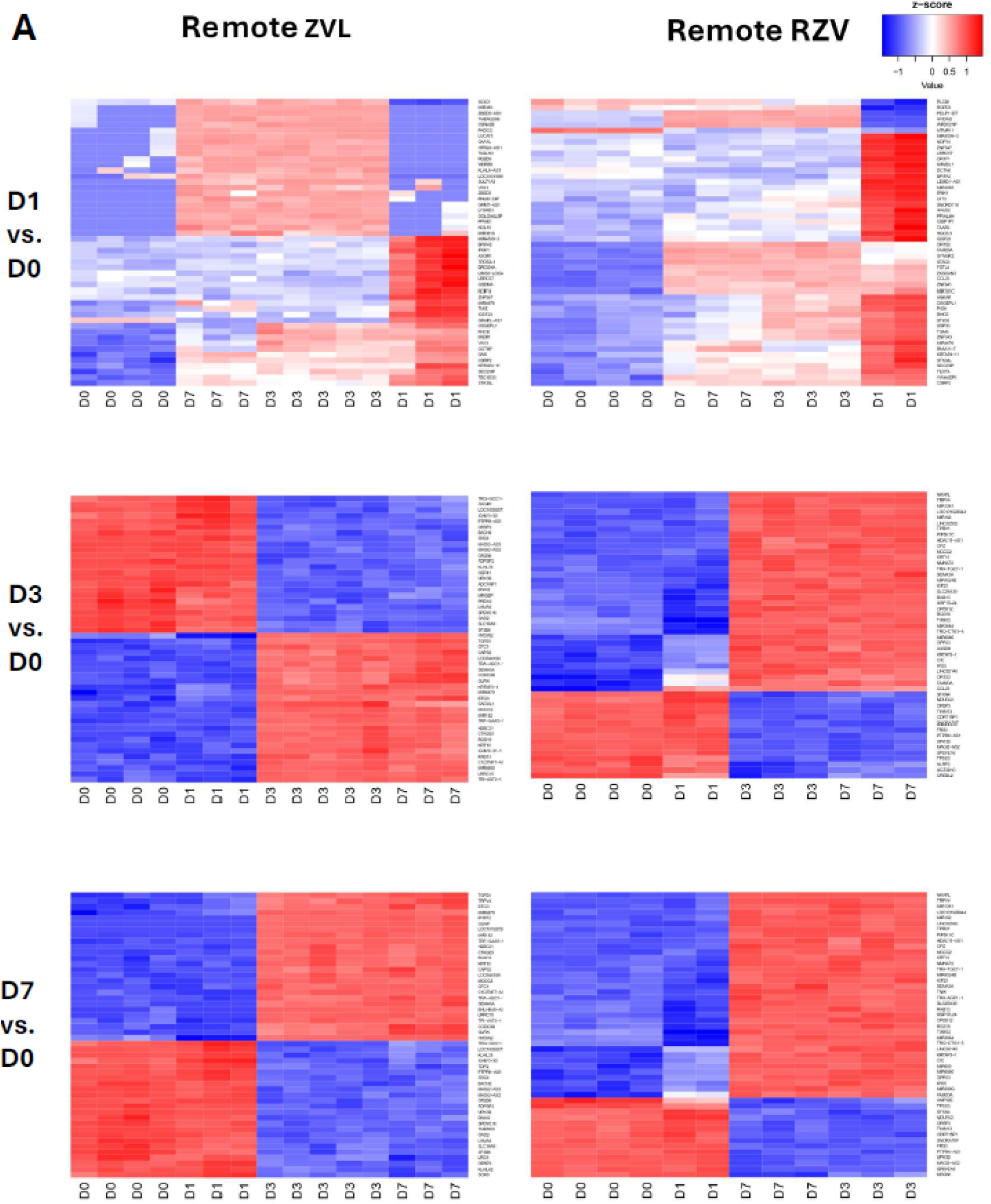
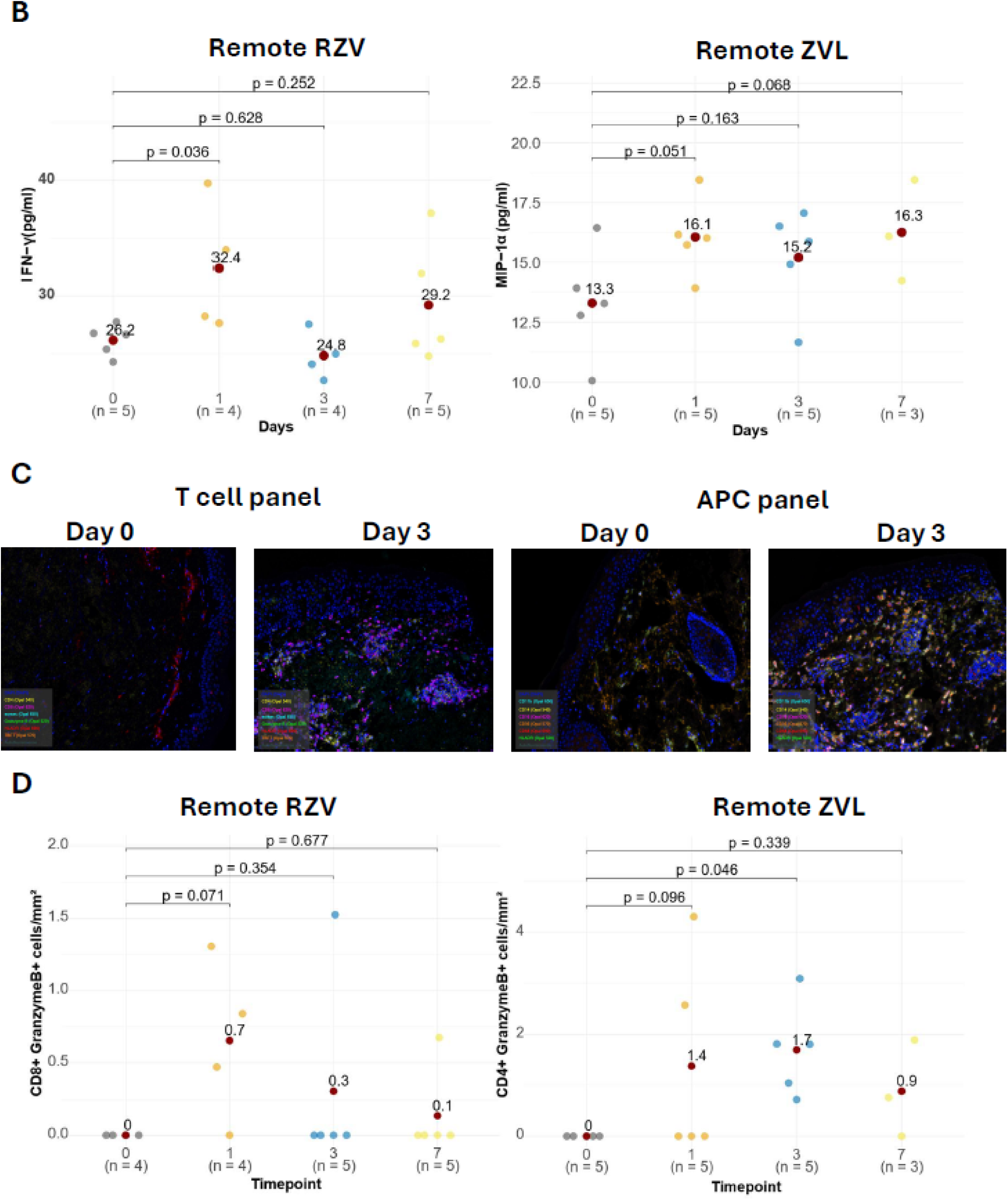
Immune responses at the cutaneous site of VZV vOka replication. Data were derived from 18 participants in the <u>Remote RZV</u> and 18 in the <u>Remote ZVL</u> groups. **Panel A** shows heatmaps of significantly differentially expressed genes (DEGs) at Days 1, 3, and 7 compared to Day 0 (FDR adjusted p<0.05). Days are grouped in unsupervised clusters based on the expression of the DEGs identified in each heatmap. **Panel B** shows examples of increases in the local cytokine production in skin homogenates. The dots represent individual participant results color-coded by timepoint. The brown dots and numeric values represent means. The number of samples contributing data at each timepoint in each group are shown on the x axis. Horizontal bars show post-hoc pairwise p values for comparisons between Day 0 and subsequent timepoints for analytes that showed significant changes over time using ANOVA with FDR-adjusted p < 0.1. **Panel C** shows typical immunofluorescence images of the immune cell infiltrates pre- and on Day 3 post-viral inoculation. **Panel D** shows typical examples of specific immune cell densities pre- and post-infection. Additional cytokines and immune cell subsets with significant differences across timepoints are shown in **Supplementary** Figure 4 (<u>Remote ZVL</u> recipients) and **Supplementary** Figure 5 (<u>Remote RZV</u> recipients).

### Local cytokine and chemokine responses to the VZV vOka challenge

Due to the small number of samples and in the interest of maximizing the sensitivity of the analysis of the cytokine, chemokine and immune cell responses in the skin biopsies, we defined significant differences in these analyses by FDR-adjusted p value < 0.1. The cytokines and chemokines measured in plasma (above) were also measured by chemiluminescence microarray in 18 tissue homogenates from the <u>Remote ZVL</u> and 18 from the <u>Remote RZV</u> groups. Increases in local IL-6, IL-12p70, IL-17A, IFN-α2a, MIP-1α, and TNF-α occurred in the <u>Remote ZVL</u> group starting on Day 1 (**Figure 6B, Supplementary Figure 4**). MIP-1α, IL-2, and IL-12p70 concentrations remained higher than pre-challenge levels up to Day 7. In contrast, <u>Remote RZV</u> participants showed limited cytokine activation exclusively represented by IFN-γ, which increased on Day 1 and approached pre-challenge levels on subsequent days (**Figure 6B**). These results were consistent with the RNAseq analysis and indicated more intense local cytokine stimulation in the <u>Remote ZVL</u> than the <u>Remote RZV</u> group. The corollary of this observation is that <u>Remote RZV</u> recipients might have controlled the viral replication faster than <u>Remote ZVL</u> recipients, limiting the local inflammation.

### Local cellular immune responses to the VZV vOka challenge

The analysis of 18 biopsies from the <u>Remote ZVL</u> and 18 from the <u>Remote RZV</u> participants showed that the density of immune cells in skin was very low before the challenge and increased by multiple orders of magnitude post-inoculation in both vaccine groups (**Figure 6C**). In the <u>Remote ZVL</u> group, most innate and adaptive immune cells reached peak densities on Day 3 compared to Day 0, including CD86+HLA-DR+ macrophages, CD14+HLA-DR+ monocytes, NK cells, total CD4+ and CD8+ T cells, CD4+ and CD8+ Tbet+ effector T cells (Teff), CD4+GrB+ CTL, and CD8+HLA-DR+ activated T cells (p≤0.07; **Figure 6C, Supplementary Figure 4**). CD4+Eomes+ memory T cells (Tmem) significantly increased on Day 7 compared to Day 0 (p=0.06; **Supplementary Figure 4**). CD8+Eomes+ Tmem also increased on Day 7 without reaching significance (**not depicted**). In the <u>Remote RZV</u> group, the most prominent changes in the cellular infiltrate occurred on Day 1, when CD8+GrB+ CTL and CD8+Tbet+ Teff densities increased compared to Day 0 (p≤0.07; **Figure 6D**; **Supplementary Figure 5)**. Total CD4+ and CD8+ T cells peaked on Day 3, but only the increase in CD4+ T cells reached significance. CD4+Eomes+ and CD8+Eomes+ Tmem increased on Day 7 without reaching significance (**not depicted**). CD14+HLA-DR+ monocytes significantly increased on Day 7 (**Supplementary Figure 5**). Overall, compared to the <u>Remote ZVL</u>, the <u>Remote RZV</u> group displayed faster migration of Teff and CTL to the site of infection and lower cellular infiltrate after Day 1.

### Clonotypic analysis of the skin T cell infiltrate

We explored the hypothesis that VZV replication in the skin led to the migration of VZV-specific T cells to the site of viral replication. To test this hypothesis, we initially selected 5 <u>Remote RZV</u> and 5 <u>Remote ZVL</u> recipients with skin biopsies on Day 3, when T cells reached maximum densities in both vaccine groups (**Figure 7A**). After reviewing the Day 3 results, we added samples from 4 <u>Remote RZV</u> recipients with Day 1 skin biopsies to the analysis. Day 1 biopsies were studied only in <u>Remote RZV</u> recipients because only this group displayed increases in CD8+ T cells at the site of inoculation as early as Day 1 post-challenge (**Supplementary Figure 6 A**). We also measured VZV-specific T cell clonotypes in PBMC from Day 0 collected from the participants whose skin biopsies were analyzed to determine if VZV-specific T cells in the circulation were available to migrate to the site of viral replication.

**Figure 7.**
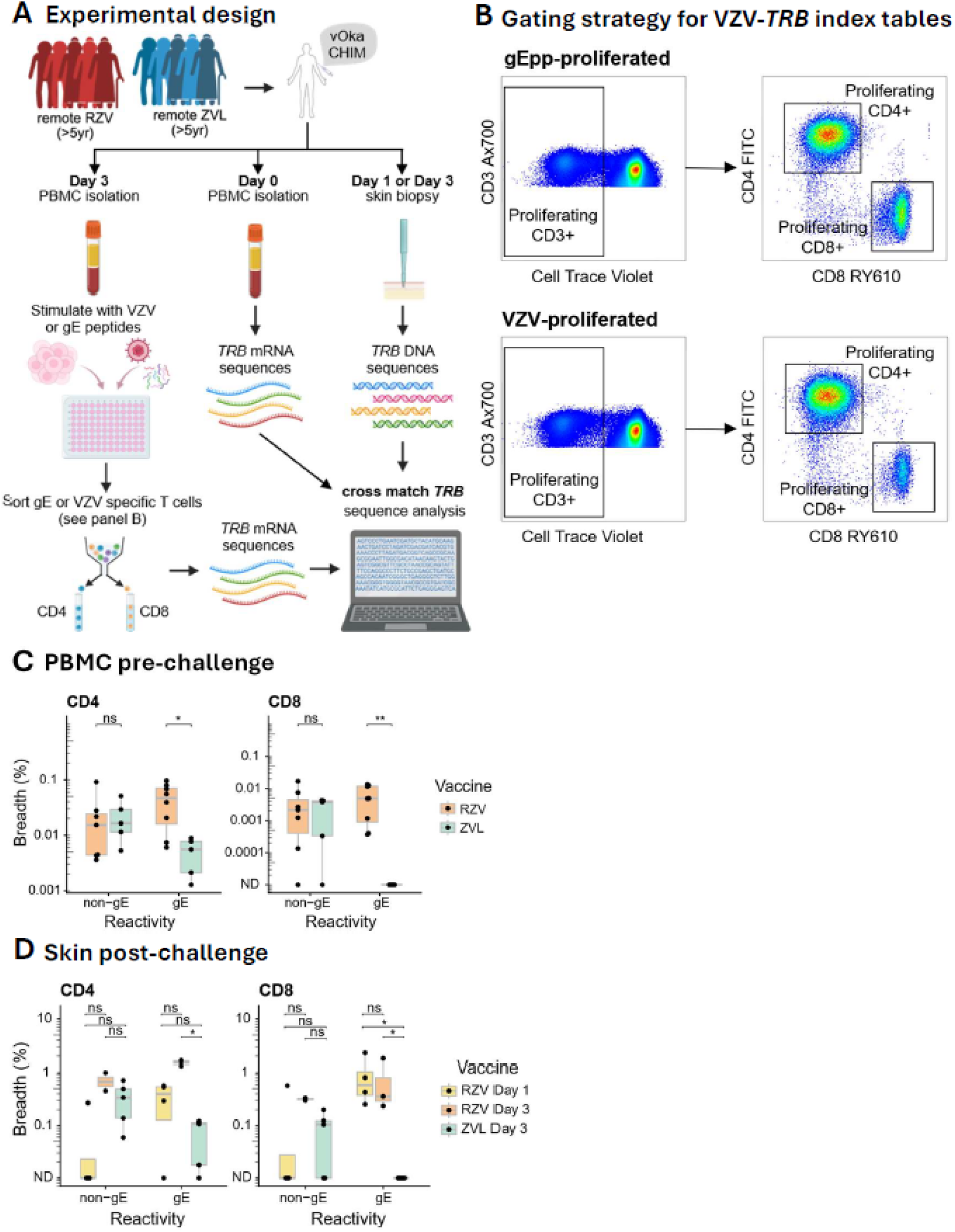
Clonotypic analysis of the T cell infiltrate in skin and relationship with pre-challenge circulating T cells in blood. Panel A shows the schematic representation of the study design. **Panel B** shows the gating strategy for sorting CD4+ and CD8+ T cells expanded in vitro with VZV-gE-peptide stimulation (top row) or VZV lysate stimulation (bottom row). **Panel C** shows the analysis of CD4+ and CD8+ VZV-gE-matched *TRB*s (expanded by VZV-gE peptide stimulation) and CD4+ and CD8+ VZV-non-gE-matched *TRB*s (exclusively expanded by VZV-lysate stimulation) in PBMC pre-challenge in 5 <u>Remote ZVL</u> and 8 <u>Remote RZV</u> participants. p values for comparisons between vaccine groups were calculated using unpaired Wilcoxon test. Asterisks represent p<0.05. **Panel D** shows the analysis of CD4+ and CD8+ VZV-gE-matched and VZV-non-gE-matched *TRB*s in skin biopsies on Day 1 in 4 <u>Remote RZV</u> participants and on Day 3 in 5 <u>Remote ZVL</u> and 2 <u>Remote RZV</u> participants with available data. p values calculated and represented as above.

We generated within-person lookup tables of VZV-reactive TCRs by stimulating each donor’s PBMC from Day 3 post-challenge with whole, inactivated VZV lysate or VZV-gE overlapping peptides. CD4+ and CD8+ T cells that proliferated in response to stimulation were sorted for TCRβ (*TRB*) sequencing as described in the Methods section (**Figure 7B** shows the gating strategy). Using the same methods for *TRB* sequencing, we characterized VZV-specific T cell clonotypes in Day 0 bulk PBMC. *TRB* in biopsies were analyzed using a genomic DNA-based method in which *TRB* read numbers were proportional to cell numbers (Adaptive Biotechnologies). *TRB* data from skin and Day 0 PBMC were compared within-person to *TRB* from sorted cells lookup tables to identify CD4+ and CD8+ clonotypes that matched VZV-gE- and/or VZV lysate epitope-reactive clonotypes expanded from blood. Matching clonotypes were defined as those sharing CDR3 amino acid sequence and V gene usage when compared within-person. Clonotypes present in either VZV-gE- alone or both VZV-gE- and VZV-lysate-expanded T cells were defined as VZV-gE-reactive. Clonotypes exclusively identified in VZV-lysate-expanded T cells were defined as VZV-non-gE-reactive.

The proportion of unique VZV-gE-reactive clonotypes out of total unique clonotypes, which defined the breadth in bulk Day 0 PBMC, was higher in <u>Remote RZV</u> than in <u>Remote ZVL</u> recipients (median 5.2 x10^-5^ vs 6.0 x10^-4^, p = 0.01; **Figure 7C**). Notably, VZV-gE-matched CD8+ clonotypes were exclusively found in <u>Remote RZV</u> recipients (**Figure 7C**). The breadth of VZV-non-gE-reactive unique clonotypes did not differ between vaccine groups (median 1.6 x10^-4^ vs 1.8 x10^-4^, p = 0.64).

Overall, Day 3 skin samples yielded 80 to 11,549 *TRB* clonotype sequences (defined by amino acid sequence, V and J gene usage) per biopsy, collapsing to 80 to 9044 unique clonotypes (by amino acid sequence) per biopsy. Two biopsies from <u>Remote RZV</u> recipients had a total of 80 and 112 unique clonotypes each after submission of several sections for sequencing. We interpreted the low number of clonotypes as insufficient sampling and excluded these biopsies from the *TRB* specificity analysis. In the three <u>Remote RZV</u> recipients with adequate sampling, we detected 662 to 5072 total *TRB* sequences, representing 629 to 4460 unique clonotypes. In the <u>Remote ZVL</u> group a total of 1759 to 11,549 total *TRB* sequences, composed of 1641 to 9044 unique clonotypes, were identified in each skin fragment. This suggests that more inflammation was detected in <u>Remote ZVL</u> compared to <u>Remote RZV</u> recipients at the site of vOka injection, however a greater proportion of the detected clonotypes in <u>Remote RZV</u> recipients matched sequences identified as gE-reactive. The median (range) proportion of unique clonotypes that matched VZV-gE-reactive CD4+ clonotypes from autologous PBMC was 1.6% (1.1-1.7%) in <u>Remote RZV</u> recipients compared to a median (range) of 0.11% (0-0.12) in <u>Remote ZVL</u> recipients (p = 0.035). In <u>Remote RZV</u> recipients, a median of 0.36% (0.2-1.2%) of unique clonotypes matched CD8+ VZV-gE reactive clonotypes, whereas in <u>Remote ZVL</u> recipients no sequences matching CD8+ VZV-gE reactive clonotypes were observed in skin (median 0%, p = 0.017). Among VZV-non-gE-reactive clonotypes, there were no differences between <u>Remote RZV</u> and <u>Remote ZVL</u> recipients in Day 3 skin CD4+ median (range) [0.3% (0-0.9%) in <u>Remote RZV</u> recipients, 0.3% (0.06-0.7%) in <u>Remote ZVL</u> recipients] or CD8+ median (range) [0.3% (0-0.3%) in <u>Remote RZV</u> recipients vs 0.07% (0-0.6%) in <u>Remote ZVL</u> recipients; (**Figure 7D**)]. Considering all sequences detected in skin, the sum of *TRB* sequences as well as CD4+ and CD8+ VZV-gE-reactive and VZV-non-gE-reactive were only slightly higher than the number of unique clonotypes, indicating that most clonotypes in skin were detected at single copy.

After uncovering the presence of VZV-gE-specific T cell clonotypes in the peripheral blood of <u>Remote RZV</u> recipients and having demonstrated that only <u>Remote RZV</u> recipients had >100 CD8+ T cells/mm^2^ of skin on Day 1 (**Supplementary Figure 6**) and significant increases in CD8+ CTL and Teff at the site of viral replication (**Figure 6D**; **Supplementary Figure 5)**, we proceeded to seek VZV-reactive T cell clonotypes in the Day 1 skin biopsies obtained from the 4 <u>Remote RZV</u> participants with available samples. The *TRB* sequencing in biopsies revealed 175 to 2366 total clonotypes, including 169 to 2109 unique clonotypes. Of these, the median (range) breadth of unique autologous-matching VZV-gE-reactive CD4+ and CD8+ clonotypes were 0.42% (0.0–0.57%) and 0.62% (0.25-2.3%), respectively, and of VZV-non-gE-reactive CD4+ and CD8+ clonotypes 0% (0-0.27%) and 0% (0-0.57%), respectively (**Figure 7D**).

Taken together, the use of blood-derived VZV- or gE-reactive autologous *TRB* sequences as within-person barcodes to measure virus-specific T cells in skin at Day 1 and Day 3 post-inoculation showed that the T cell infiltrate largely reflected the hierarchy of VZV-specific T cell clonotypes in Day 0 PBMC. RZV was associated with availability of VZV-gE-specific T cells in blood and a larger and faster VZV-specific T cell response at the site of viral replication. Interestingly, gE-reactive CD8+ T cell clonotypes identified in blood after RZV, but before challenge, were also detected in the skin shortly after the VZV vOka challenge.

## Discussion

We developed a CHIM for VZV reactivation using intradermal inoculation of replication-competent VZV vOka vaccine strain. We validated that vOka CHIM mimicked wild-type VZV reactivation by showing that vOka expressed VZV transcripts consistent with viral replication at the site of inoculation. The cellular infiltrate at the site of the vOka challenge was similar to the infiltrate previously described in ganglia experiencing VZV reactivation during HZ ^26^. The vOka inoculation also resulted in VZV DNAemia as described in patients with HZ or with asymptomatic VZV reactivations ^14^. Moreover, the model clarified the significant differences between recent and remote HZ vaccine recipients. These properties of the model allowed us to dissect the local and systemic immune responses to VZV replication, to identify immune correlates of control of viral replication, and to study the differential effect of ZVL and RZV on the immune responses to VZV replication.

Our novel findings contribute to understanding the superior protective effect of RZV compared to ZVL against HZ. RZV recipients have VZV-gE-reactive CD4+ and CD8+ T cells circulating in the blood of RZV recipients for ≥5 years post-vaccination. In contrast, ZVL recipients do not have detectable VZV-gE-matched CD8+ T cells in blood or at the site of viral replication and have much fewer VZV-gE-reactive CD4+ T cells than RZV recipients both in blood and at the site of viral replication. The corollary of these observations is that VZV-gE-matched T cells in RZV recipients are uniquely generated by this vaccine. While gE is the most abundant glycoprotein in VZV and ZVL has been shown to stimulate VZV-gE-specific immunity^27^, the context for VZV-gE antigen presentation in RZV and specifically the AS01B adjuvant are distinct from that after subcutaneous ZVL, likely accounting for the superior immune response to VZV-gE with RZV. In addition, the Remote ZVL group had a delayed local CD8+ T cell response post-infection compared to RZV. The persistence of VZV-gE-reactive T cells in blood facilitates a more robust and rapid response to VZV replication by RZV than ZVL recipients. This observation was particularly evident with respect to CD8+ CTL, which increased on Day 1 post-inoculation in RZV recipients, but did not significantly increase in ZVL recipients during the entire period of observation. Previous studies showed that ZVL administration induced relatively high VZV-gE-specific CD4+ T cell responses, but fewer VZV-gE-specific CD8+ T cells ^28^. Likewise, in the context of HZ, members of our group found high abundance of VZV-gE-reactive CD4+ T cell clonotypes and low abundance of CD8+ T cell clonotypes in skin ^29^. These results address two subjects of debate in the medical literature: (1) we show that the early T cell infiltrate at the site of viral replication contains T cells specific to cognate antigens in vaccine recipients as opposed to nonspecific, innate-like CD8+ T cells as previously suggested ^30^; and (2) we show VZV-gE-reactive CD8+ T cells exclusively in the blood and skin of RZV recipients, suggesting that they were generated or expanded by the vaccine, which supports our previous findings ^31^ and differs from other reports ^32^. Notably, a COVID-19 adjuvanted protein vaccine also induced CD8+ T cell responses in humans ^33^ challenging a long-held notion that protein vaccines do not stimulate CD8+ T cell responses.

We found VZV-non-gE-specific CD4+ and CD8+ T cells in blood before the VZV challenge both in RZV and ZVL recipients. In ZVL recipients, these cells may have resulted from the response to vaccination. However, it is more likely that these cells were maintained in circulation since primary infection through a mechanism independent of vaccination, considering that VZV-specific CMI in blood returns to pre-vaccination levels 2 years post-ZVL in >90% of vaccinees ^34^. Alternatively, asymptomatic VZV reactivations, which are not uncommon in older adults ^15^, may periodically boost VZV-CMI in blood of both ZVL and RZV recipients.

The rapid and robust early immune response to replicating VZV in <u>Remote RZV</u> recipients likely decreased viral replication compared with <u>Remote ZVL</u> recipients. This conclusion was derived from the higher antibody response to the viral challenge in <u>Remote ZVL</u> than <u>Remote RZV</u> recipients, which is typically proportional to the amount of antigen generated by viral propagation ^23,25^ and from the higher local and systemic inflammation in <u>Remote ZVL</u> compared to <u>Remote RZV</u> recipients. The symptoms that characterize HZ result from inflammation and neuronal destruction in the sensory ganglia, where VZV reactivates, and in the skin, where the virus migrates after reactivation. Thus, decreased inflammation at the site of VZV reactivation may play an important role in the RZV-mediated protection against clinically evident HZ and in the attenuation of symptoms and lower incidence of complications in breakthrough cases ^19^. Overall, the superior efficacy of RZV likely relies on the rapid mobilization of these protective immune responses after VZV reactivation. RZV is unlikely to prevent VZV reactivation, because of the following: (1) VZV-gE is not transcribed or translated during latency ^35^ and (2) VZV-infected dorsal root ganglia generally do not show appreciable levels of T cell accumulation during latency, with a single example in the literature of the recovery of a VZV-reactive, HSV non-cross-reactive, CD8+ T cell clonotype from a VZV-infected, HSV non--infected trigeminal ganglion at autopsy ^36,37^. Our findings indicate that the memory immune cells induced by RZV persist in circulation for at least 5 years ensuring rapid suppression of reactivated VZV and preempt the florid inflammatory response that accompanies HZ. Identifying and characterizing the molecular mechanisms that underly the persistence of the VZV-gE-specific T cells in circulation is essential for the development of additional effective vaccines in older adults.

We found VZV DNA in blood and VZV mRNA in the skin up to 7 days post-inoculation in the CHIM. These results are in accordance with our previous findings of VZV DNAemia for up to 14 days in people vaccinated with ZVL for the first time ^24^. Here, we used VZV DNAemia on Day 3 post-challenge as a surrogate marker of viral replication after suspecting that DNAemia on Day 1 might have included viral DNA from the inoculum. We determined that the frequency of CD4+ T cells with cytotoxic characteristics in blood was the primary immune correlate with control of viral replication. This finding is in agreement with previous reports demonstrating that CD4+ CTL play an important role in clearing herpesvirus infections ^38^. We and others previously showed that high frequencies of VZV-specific CD8+ CTL in blood were associated with protection against HZ in immunocompromised hosts ^25,39,40^. This CHIM showed that VZV-gE- and non-gE-specific CD8+ CTL migrate to the site of viral replication in RZV recipients on Day 1 post-inoculation, which likely contributed to the control of infection. In ZVL recipients, we found an increase in CD4+ CTL at the site of viral replication delayed until Day 3 post-inoculation. Collectively, these data suggest that both CD4+ and CD8+ CTL contribute to the control of VZV replication and may be essential components of protective vaccine responses.

The differences we found in the systemic immune responses of the two vaccines after the viral challenge were most prominent between the <u>Remote ZVL</u> and <u>Recent RZV</u> groups, which were likely accentuated by the time since vaccination. It is known that the efficacy of ZVL rapidly wanes over time and is <30% at 5 years after vaccination ^41^, whereas the efficacy of RZV remains >80% ^19^. Thus, we expected the largest immune response differences in this CHIM between the two <u>Remote</u> groups. Moreover, we <u>found</u> very few significant immune response differences between <u>Recent RZV</u> and <u>Remote RZV</u> recipients, consistent with the durability of the efficacy of this RZV. In addition, we also found fewer differences between the <u>Recent RZV</u> and <u>Recent ZVL</u> groups than between the <u>Recent RZV</u> and <u>Remote ZVL</u> groups. Notably, the <u>Recent ZVL</u> recipients were significantly younger than the other study participants and it is well known that age at the time of vaccination has a prominent effect on the efficacy of ZVL^16^. Taken together, these data support the utility of the CHIM that we developed for studying protective immune responses against HZ.

As mentioned above, our study had limitations imposed by the relatively small number of skin biopsies. Most of the participants were white non-Hispanic, which may influence the generalizability of our findings. There was an age difference between the <u>Recen</u>t and <u>Remote</u> vaccine recipients with <u>Recent</u> vaccinees being younger, which was inherent to the study design. T cell clonotype matching used only *TRB* sequences rather than dual-chain TCR data, and it is possible that some T cells proliferating in response to gE peptide pools or whole VZV antigen were cytokine-responsive rather than recognizing antigen via specific TCR. Lastly, we used DNAemia as a marker of viral replication, which does not equal viremia, although it is widely accepted as a surrogate ^14^.

We conclude that this CHIM of VZV reactivation indicates the primary role of CTL in protection against VZV replication. We were also able to define certain responses to VZV replication that differentiate the highly efficacious RZV from the less efficacious ZVL, including the persistence of VZV-gE-specific circulating CD4+ and CD8+ T cells, rapid and robust VZV-gE-reactive CD8+ T cell infiltration at the site of viral replication, and limited local inflammation. Importantly, we demonstrated that VZV-specific T cells infiltrate the site of viral replication as early as Day 1 post-infection in skin and presumably in ganglia emphasizing the protective role of circulating antigen-specific T cells. These findings underscore desirable characteristics for new vaccines against herpesviruses in adults.

## Methods

### Study design and participants

This study (NCT 04169009) was approved by the Colorado Multiple Institutions Review Board. All participants signed informed consent. Adults ≥ 50 years of age, without any immunocompromising conditions were enrolled in 4 groups: Remote ZVL and Remote RZV groups, who received ZVL or RZV, respectively, ≥ 5 years before receiving a vOka challenge in the form of Zostavax™ intradermal on Day 0; and Recent ZVL and Recent RZV groups, who were immunized at enrollment and challenged with vOka 5 to 12 months after immunization. The intradermal injection was performed as previously described ^22^. All participants had blood collected on Day 0 before the challenge and on Days 1, 3, and 7 post-challenge. Consenting participants in the Remote vaccine groups also had punch skin biopsies performed on Days 0, 1, 3, and 7 at the intradermal challenge site in groups of 3 to 5 independent participants per group per Day.

### Specimen processing

Peripheral blood mononuclear cells (PBMCs) were isolated from heparinized whole blood by density-gradient centrifugation using Ficoll-Hypaque (Sigma-Aldrich) and cryopreserved as previously described^42^. Plasma was collected following centrifugation of whole blood, aliquoted to minimize freeze–thaw cycles, and stored at −80°C until analysis.

Skin biopsies were collected and transported to the laboratory within 2h of procurement. Each biopsy specimen was divided into two equal portions. One half was fixed in 10% neutral-buffered formaldehyde for 24h, washed with PBS (Corning), and transferred to 70% ethanol for histological processing.

The remaining half of the biopsy was suspended in 600 µL of RLT Plus buffer (Qiagen) and mechanically homogenized using the Qiagen TissueLyser LT according to the manufacturer’s instructions. RNA was extracted from 480 µL of tissue homogenate using the RNeasy Plus Mini Kit (Qiagen) per the manufacturer’s protocol and stored at -80°C until used in sequencing assays. An additional 120 µL of the homogenate was aliquoted and stored at −80°C for cytokine and chemokine analysis.

### RNA sequencing from skin homogenates

The concentration of RNA extracted as above concentration was assessed using a NanoDrop spectrophotometer (Thermo Fisher Scientific) and quality was assessed using the Agilent TapeStation High Sensitivity RNA ScreenTape assay. RNA integrity was evaluated using the RNA Integrity Number equivalent (RINe). When a RINe score could not be assigned, the RNA integrity was below the assay’s recommended range and samples were excluded from further analysis. RNAseq libraries were prepared from 200 ng of total RNA using the Universal Plus mRNA-Seq Library Preparation Kit with NuQuant (Tecan) according to the manufacturer’s instructions. Libraries were sequenced on an Illumina NovaSeq 6000 platform to generate 150-bp paired-end reads at a target depth of 80 million paired-end reads per sample. Raw sequencing reads were demultiplexed using bcl2fastq.

### Immune cell infiltrate characterization by immunofluorescence

Through our collaboration with the Human Immune Monitoring Shared Resource (HIMSR) at the University of Colorado School of Medicine we performed 7-color multispectral imaging using the Vectra 3 instrument (Akoya Biosciences). To analyze tissue infiltrate in formalin-fixed paraffin-imbedded biopsy sections, the tissues were sequentially stained with Granzyme B, CD4, Tbet, EOMES, CD8, and HLADR (Resources Table) or CD16, CD68, CD56, CD11b, HLADR, and CD14 (Resources Table). Briefly, the slides were deparaffinized, heat treated in antigen retrieval buffer, blocked, and incubated with primary antibody, followed by horseradish peroxidase (HRP)-conjugated secondary antibody polymer (Akoya Biosciences), and HRP-reactive OPAL fluorescent reagents (Akoya Biosciences) on a Bond RX autostainer (Leica Biosystems). To prevent further deposition of fluorescent dyes in subsequent staining steps, the slides were stripped between each stain with heat treatment in antigen retrieval buffer. Slides were stained with spectral DAPI prior to mounting coverslips with aqueous mounting media. Whole slide scans were collected using the 10x objective and multispectral images were imaged using the 20x objective with a 0.5 micron resolution. The 7 color images were analyzed with inForm software to unmix adjacent fluorochromes, subtract autofluorescence, segment the tissue, compare the frequency and location of cells, segment cellular membrane, cytoplasm, and nuclear regions, and phenotype infiltrating immune cells.

### Phenotypic characterization by flow cytometry

Cryopreserved PBMCs were thawed, washed with PBS, and plated at 5 × 10 cells per well in 96-well plates before staining. Two multicolor flow cytometry panels were used to characterize antigen-presenting cells (Panel 1) and T cells and natural killer (NK) cells (Panel 2).

Panel 1 – Antigen-Presenting Cells: PBMC were incubated with Zombie Aqua viability dye (Biolegend) and then washed in stain buffer (1% BSA in PBS; Sigma-Aldrich, Corning). Cells were surface stained in brilliant stain buffer plus (BD Biosciences) with the following antibodies: CD14 BV605 (BioLegend), CD123 BV785 (BioLegend), CD16 FITC (BioLegend), CD40 PerCP-Cy5.5 (BioLegend), CD1c PE-CF594 (BioLegend), PDL1 PE-Cy7 (BioLegend), CD141 APC (BioLegend), CD3 Ax700 (BD Biosciences), CD19 Ax700 (BioLegend), CD20 Ax700 (BioLegend), CD56 Ax700 (BioLegend), and HLA-DR APC-H7 (BD Biosciences). True-stain monocyte blocker (Biolegend) was also included. After washing, cells were fixed and permeabilized using BD FACS Lysing Solution and BD FACS Permeabilizing Solution 2, followed by intracellular staining with TNFα PE (BD Biosciences) and IL-10 BV421 (BioLegend).

Panel 2 – NK and T Cells: PBMC were incubated with Brefeldin A (Sigma, 5 µg/mL), Monensin (Sigma, 5 µg/mL), and anti-CD107a PerCPCy5.5 (BD Biosciences) for 4h before staining. Cells were washed with PBS, stained with Zombie Aqua viability stain (Biolegend), and washed in stain buffer before surface staining in Brilliant Stain Buffer. Surface antibodies included CD95 BV421 (BioLegend), CD4 BV570 (BioLegend), CD38 BV650 (BioLegend), CD16 BV785 (BioLegend), CD56 PE (BioLegend), CCR7 PE-D594 (BioLegend), HLA-DR APC (BD Biosciences), CD3 Ax700 (BD Biosciences), and CD45RO APC-H7 (BD Biosciences). True-stain monocyte blocker was also included. After washing, cells were fixed and permeabilized as above and intracellularly stained with Bcl2 Ax488 (BioLegend), IFN-γ PE-Cy7 (BioLegend), and Ki67 BV605 (BioLegend).

For both panels, cells were washed and fixed with PBS+1% paraformaldehyde before acquisition on the Novocyte Quanteon flow cytometer (Agilent). Analysis was done using FlowJo v10 (BD Biosciences) and the gating strategy is shown in **Supplementary Figure 1**. 1.

#### VZV IFNγ ELISPOT

Cryopreserved PBMC were thawed and rested overnight at 37°C and 5% CO_2_ at 10^6^ PBMC/mL in growth medium consisting of RPMI 1640 with L-glutamine (Gemini BioProducts), 10% human AB serum (Gemini BioProducts), 2% HEPES buffer (Mediatech), and 1% penicillin-streptomycin (Gemini BioProducts). The next day, 96-well Millipore ELISpot plates (Sigma) were coated with anti-human IFNγ antibody (Mabtech, mAb1-D1k). Rested PBMC were added at 250,000 cells/well in duplicate wells and stimulated for 48 h at 37°C, in a humidified atmosphere and 5% CO_2_ with UV-irradiated VZV-infected cell lysate, mock-infected control, and PHA. A study-specific leucopack PBMC sample with known VZV reactivity was included in each run for assay validation. Plates were incubated with secondary anti-human IFN conjugated with (Mabtech, mAb 7-B6-1) and developed with the colorimetric substrate 5-Bromo-4-chloro-3-indolyl phosphate/ nitro blue tetrazolium (Mabtech). Results were reported as the mean number of spot-forming cells (SFC)/10^6^ PBMC in VZV-stimulated wells after subtraction of SFC in mock-stimulated wells. Results were considered valid if the leucopack control tested within pre-specified upper and lower limits, if the number of SFC/10^6^ PBMC in antigen-stimulated wells was ≥2-fold higher than in mock-stimulated wells and if PHA-stimulated wells had ≥ 400 SFC/10^6^ PBMC.

#### Anti-VZV antibodies

were measured by quantitative ELISA using the VZV IgG ELISA kit from Gold Standard Diagnostics Corp (Davis CA, GSD01-180) as per manufacturer’s instructions. The antibody concentration was calculated by interpolation onto the control standard curve provided by the manufacturer and reported in ELISA units (EU)/mL.

#### Cytokine analysis in blood and tissue

Frozen plasma and biopsy supernatants were thawed tested using the Meso Scale Discovery U-PLEX custom human biomarker assays, which included GM-CSF, IFN-, IL-2, IL-6, IL-10, IL-12p70, IL-17A, TNF-α, IFN-α2a, IL-15, including MCP-1, MIP-1α, and MIP-1β. Assays were performed per kit protocol, read on a MESO QuickPlex SQ120 instrument, and concentrations determined with MSD DISCOVERY WORKBENCH 4 analysis software.

#### VZV DNA PCR in blood and tissue

DNA was extracted from 200ul of plasma and buffy coat with QIAamp DNA Blood Mini kit (Qiagen, Cat# 51104) and 20µm FFPE Tissue sections with QIAamp DNA FFPE Tissue Kit (Cat# 56404). 10ul of extracted DNA were added to 10ul of VZV DNA PCR master mix containing 2ul of LightCycler Fastart DNA Master Hybprobe (Roche, Cat# 12239272001), 3.5 mM of MgCl2, 0.4 µM of VZV orf63A gene primer F (5’-CGCGTTTTGTACTCCGGG-3’) and 0.4 µM of primer R (5’-ACGGTTGATGTCCTCAACGAG-3’) and 0.15 µM of TaqMan probe (5’-6FAM-TGGGAGATCCACCCGGCCAG-BBQ-3’). Reactions were carried out in the LightCycler 2.0 (Roche) in the following conditions: denaturation at 95° C for 10 min, 50 cycles of amplification for 10 s at 95° C and 30 s at 60° C. and cooling to 40° C for 30 min. This method has a limit of detection of 200 DNA copies/mL. Negative and extracted positive controls and the quantitative standard were included in each run. Results were considered valid if the controls performed within pre-established ranges.

### *TRB* analyses

#### VZV-gE- and VZV-whole virion-specific CD4+ and CD8+ T cell isolation and *TRB* sequencing

Thawed PBMCs from Day 3 or Day 7 post-challenge were labeled with CellTrace Violet (BioLegend) and cultured for 5 days in the presence of VZV UV inactivated whole virion or VZV-gE peptide pools (Genscript). Following incubation, PBMCs were washed with PBS and stained with Zombie Aqua Viability Stain (BioLegend). Cells were then surface-stained with CD3 Ax700 (BD Biosciences), CD4 FITC (Biolegend), and CD8 RY610 (BD Biosciences). Proliferating (Cell Trace Violet^dim^) CD4⁺ and CD8⁺ T cells were identified and sorted into collection buffer (1mM EDTA, 25mM HEPES, 1%BSA in PBS; Corning, Corning, Sigma-Aldrich, Corning) using the MoFlo Astrios Cell Sorter (Beckman Coulter). Sorted cells were centrifuged, supernatant was aspirated, and cell pellets were flash frozen and stored until *TRB* sequencing analysis. **Figure 7B** shows an example of the gating strategy. On the day of *TRB* sequencing, pelleted VZV-gE- and VZV-whole virion-expanded CD4+ and CD8+ T cells were submitted to RNA extraction using Qiagen Rneasy Plus Micro Kit following the manual. Extracted RNA was eluted in 12 ul RNAse-free water. 9.5 ul of each RNA preparation were used as input for *TRB* libraries build using TAKARA SMART-Seq® Human TCR (with UMIs) and related Unique Dual Index kit for further Illumina sequencing, which was performed using Illumina Nextseq 300 cycles kit with 2 to 3 million reads for each library. FASTQ files were assembled using Mixcr v4.7.0 ^43^ with Java v11.0.20 and GNU parallel 11.3.0 ^44^. Custom code used in downstream TCR sequence analysis is available for review at www.github.com/esford3/weinberg_2026.

*TRB* sequences were considered as VZV-reactive if they were present in one or both of the stimulated fractions at >1 copy, single detection clonotypes were excluded. Sequences were also excluded from the VZV-reactive reference set if they were present in the CMV external reference set ^45^. Sequences observed in the stimulated fractions from both the gE peptide and whole antigen were considered to be VZV-gE-reactive, whereas sequences only observed after stimulation with whole antigen were considered to be VZV-non-gE reactive.

#### *TRB* sequencing of PBMC

PBMC cryopreserved on Day 0 pre-challenge were thawed, counted, and 5 to 10 million cells were submitted to total RNA extraction as above. For each library build, 1ug of total RNA resuspended in less than 9.5 ul volume and processed as above. 20 million total reads in Illumina sequencing were assigned for each PBMC based library.

*TRB* sequencing of FFP-fixed biopsy tissue was performed at Adaptive Biotechnologies using the ImmunoSEQ Assay TCRBv4b.

#### Sequence-similarity analysis

Data were exported from the Adaptive Immunoseq and from the NextgenSeq platforms and analyzed in R version 4.0.1 and python 3.7. Out of frame or non-coding sequences and those without valid V or J gene assignments were excluded. Within person, unique clonotypes were defined by sequences with the same CDR3 amino acid sequence, V and J gene usage. For exact clonotype tracking (and due to the need to make comparisons across different sequencing technologies), clonotypes were considered to be present in multiple samples if they had the same CDR3 amino acid sequence and *TRBV* family (within person). Clonotypes observed in both CD4 and CD8 stimulated fractions were assigned cell type based on the fraction with higher frequency (dual CD4/CD8 assignments were not allowed).

### Statistical methods

#### Analysis of RNA-seq data

Raw sequencing data were uploaded to Galaxy platform (usegalaxy.org, analysis performed in May 2023) to process ^46^. Within the platform, the quality of raw sequencing reads was assessed using FastQC (v0.11.9). We employed Trim Galore (v0.6.7) for adapter trimming. Reads were aligned to the human reference genome (GRCh38) using Bowtie2 (v2.5.0). Gene-level read counts were quantified from the aligned BAM files using featureCounts (v2.0.8). A gene-level count matrix for all samples was downloaded and used for downstream statistical analyses in R version 4.5.1. We removed lowly expressed genes, which had fewer than 10 counts in more than 50% of the samples, resulting in 16,327 genes for remote ZVL group and 13,157 genes for remote RZV group. Differential expression analysis was performed using the DESeq2 package ^47^, which models gene counts with a negative binomial distribution. Multiple pairwise contrasts between timepoints (D1 vs. D0, D3 vs. D0, D7 vs. D0) were specified and p-values were adjusted for multiple testing using the Benjamini-Hochberg procedure ^48^. Differentially expressed genes were ranked by the adjusted p-values and absolute log2 fold change. The top ranked genes were used to generate heatmaps to visualize the expression patterns across samples.

#### VZV transcript detection and quantification

Reads that did not map to the human genome in the previous step were extracted and subsequently aligned to the VZV reference genome (NCBI Reference Sequence: NC_001348.1) using Bowtie2 (v2.5.0)^49^. Reads mapping to viral genes were counted to quantify viral transcript abundance. This approach allowed us to specifically capture viral transcripts while minimizing the bias caused by the large size difference between the viral and human genomes.

#### Immune assay data: flow cytometry, immunofluorescence, cytokines and chemokines

For tissue and cross-group comparisons, since samples were collected from independent donors, comparisons were performed using ANOVA, followed by post-hoc pairwise comparisons with p-values adjusted using the Benjamini–Hochberg procedure. This adjustment controlled for multiple comparisons across groups or timepoints for each marker separately but not applied across the entire panel of markers. Data measured in blood samples repeatedly at multiple timepoints were analyzed using repeated measures ANOVA, with timepoints as the within-subject factor and vaccine groups as the between-subject factor. Post-hoc pairwise comparisons were performed for each marker, and p-values were adjusted for multiple contrasts within that marker using the Benjamini–Hochberg procedure. No correction was applied across the entire panel of markers.

#### VZV DNA PCR

At each timepoint, samples were classified as PCR-positive or PCR-negative. PCR positivity rates of the first two groups (<u>Remote ZVL</u> and <u>Remote RZV</u>) were compared with the last two groups (<u>Recent ZVL</u> and <u>Recent RZV</u>) using chi-square test for differences in proportions. No adjustment was done across timepoints.

#### VZV DNAemia on Day 3

Given the large number of features relative to the sample size, we used a two-stage modeling approach to evaluate associations between assay data features and VZV DNAemia at Day 3. In the first stage, LASSO logistic regression ^50^ was used to select features associated with PCR positivity within each assay. Features with non-zero coefficients were then modeled jointly in a multivariable logistic regression model to assess their independent associations with DNAemia.

#### VZV antibodies by ELISA and IFNg ELISPOT

Change in antibody concentration and IFNg SFC/10^6^ PBMC from pre- to 7 days post-challenge was computed for each sample. Changes were log10-transformed after adding a small constant to ensure all values were positive prior to transformation. The transformed measures were compared across groups using ANOVA. Post-hoc pairwise group comparisons were performed, and p-values were adjusted for multiple contrast using the Benjamini–Hochberg procedure. We also fit a linear regression model on ELISPOT data to adjust for age. However, age was not significant, so we reported the unadjusted results.

## Acknowledgments

The authors thank Mss. Nancy Lang, RN and Tori Rutherford, RN for clinical and regulatory assistance, and Mr. Zelalem Segano for administrative assistance.

## Author contributions

**M.J.J.** designed experiments, coordinated study operations and project activities, collected and processed specimens, performed flow cytometry assays, analyzed and interpreted data, prepared figures, participated in manuscript preparation. **T.V.** performed statistical and bioinformatic analyses, prepared figures, interpreted data, participated in manuscript preparation. **E.S.F.** performed T-cell receptor (TCR) and bioinformatic analyses, prepared figures, and participated in manuscript writing. **L.J.** performed TCR studies and analyses and participated in manuscript preparation. **K.J.** oversaw tissue immune-cell phenotyping and analysis and participated in manuscript preparation. **M.B.** organized skin biopsy acquisition. **S.L.** performed VZV PCR assays. **T.S.** performed tissue immune-cell phenotyping and analysis. **M.C.** collected and processed specimens, performed flow cytometry assays. **J.C.** collected and processed specimens and contributed to cytokine assays. **A.B.** collected and processed specimens, performed cytokine assays, and prepared figures. **N.W.** performed VZV PCR assays. **K.J.L.** advised on TCR studies, prepared figures and participated in manuscript preparation. **S.F.** advised on the VZV transcriptome analysis. **D.M.K.** developed the strategy for TCR characterization, interpreted data, and participated in manuscript preparation. **M.J.L.** contributed to study conceptualization and CHIM design, supervised participant enrollment and study intervention, acquired funding, contributed to writing and editing the manuscript. **A.W.** conceptualized and designed the study, including the CHIM, developed methodology, administered the project, supervised the research, acquired funding, interpreted data, wrote the manuscript. All authors approved the final manuscript.

## Data Availability

RNA-sequencing data generated in this study have been submitted to the NCBI GEO, the accession number is pending. Custom code and data used for downstream T-cell receptor sequence analysis is publicly available at https://github.com/esford3/weinberg_2026. The remaining data generated and analyzed during this study are available from the corresponding author upon reasonable request.

