## Supplementary Figures for "Experimental model of VZV reactivation reveals vaccine-induced mechanisms of protection against herpes zoster": Experimental model_supplementary_figures_with_legends aw.pdf

**A**

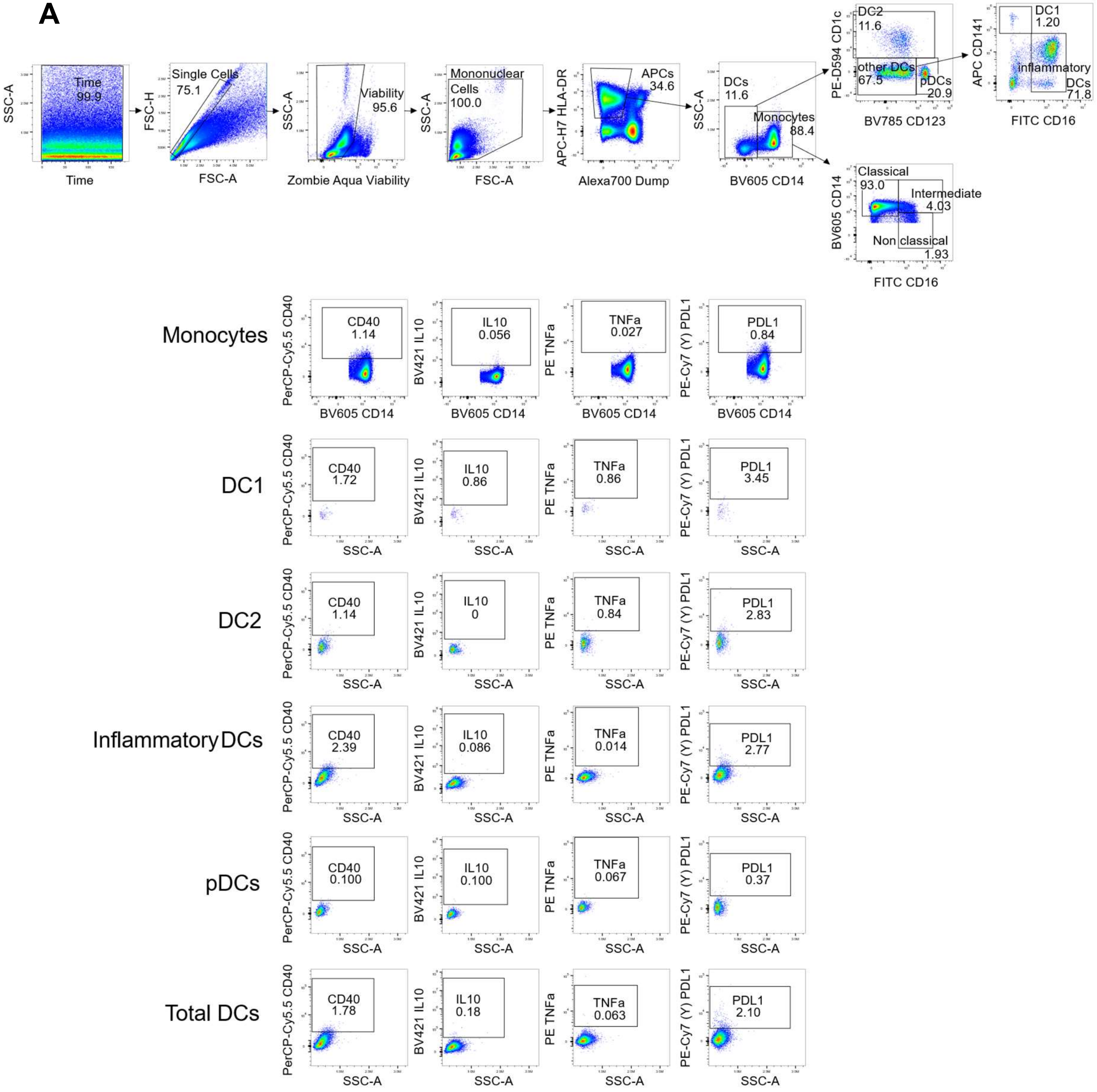

SSC-A
Time
Time 99.9
FSC-H
FSC-A
Single Cells 75.1
SSC-A
Zombie Aqua Viability
Viability 95.6
SSC-A
FSC-A
Mononuclear Cells 100.0
APC-H7 HLA-DR
Alexa700 Dump
APCs 34.6
SSC-A
BV605 CD14
DCs 11.6
Monocytes 88.4
PE-D594 CD1c
BV785 CD123
DC2 11.6
other DCs 67.5
pDCs 20.9
APC CD141
FITC CD16
DC1 1.20
inflammatory DCs 71.8
BV605 CD14
FITC CD16
Classical 93.0
Intermediate 4.03
Non classical 1.93
Monocytes
DC1
DC2
InflammatoryDCs
pDCs
Total DCs
PerCP-Cy5.5 CD40
BV421 IL10
PE TNFa
PE-Cy7 (Y) PDL1
BV605 CD14
SSC-A
CD40 1.14
IL10 0.056
TNFa 0.027
PDL1 0.84
CD40 1.72
IL10 0.86
TNFa 0.86
PDL1 3.45
CD40 1.14
IL10 0
TNFa 0.84
PDL1 2.83
CD40 2.39
IL10 0.086
TNFa 0.014
PDL1 2.77
CD40 0.100
IL10 0.100
TNFa 0.067
PDL1 0.37
CD40 1.78
IL10 0.18
TNFa 0.063
PDL1 2.10

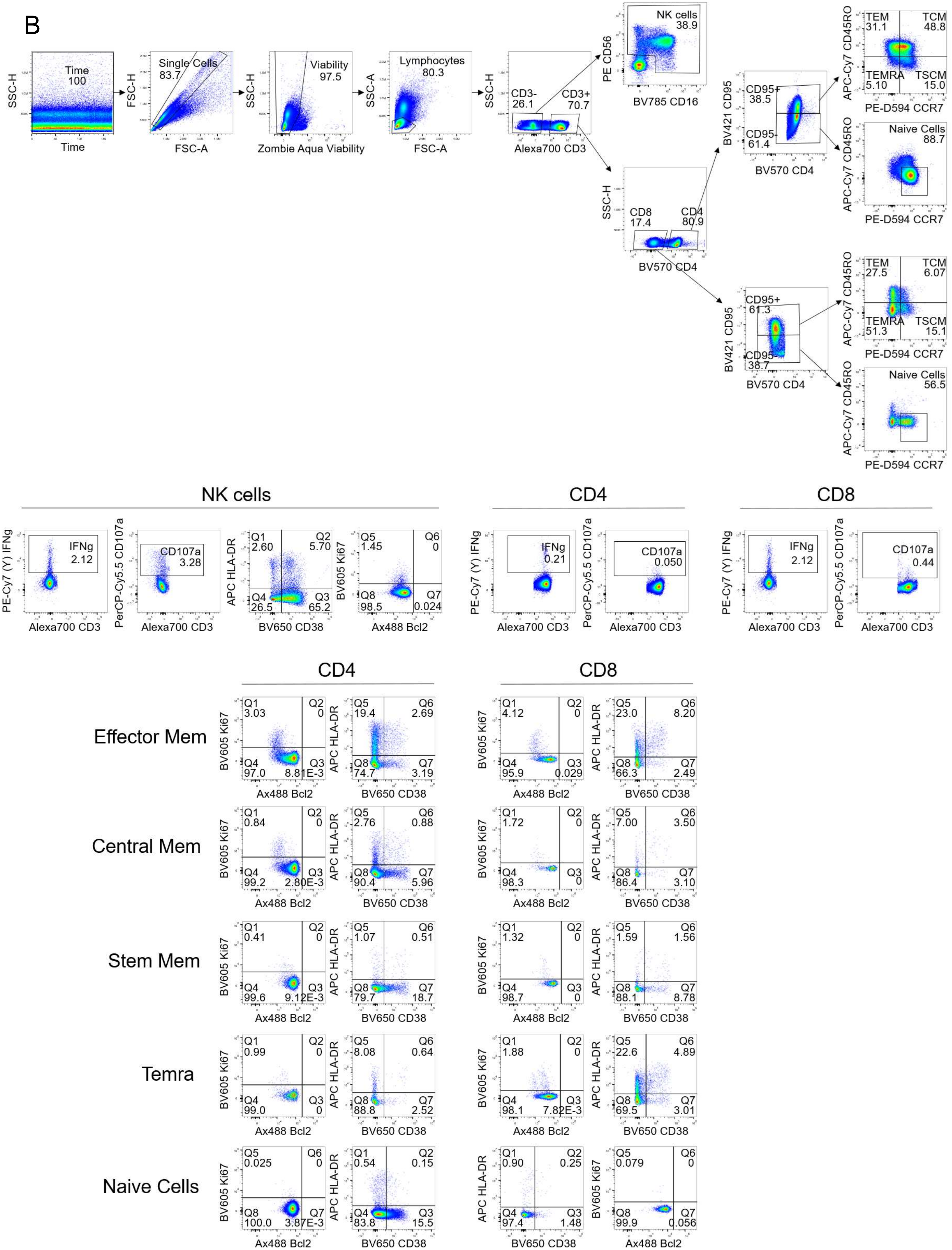
B
Time
100
SSC-H
Time
Single Cells
83.7
FSC-H
FSC-A
Viability
97.5
SSC-H
Zombie Aqua Viability
Lymphocytes
80.3
SSC-A
FSC-A
CD3-
26.1
CD3+
70.7
SSC-H
Alexa700 CD3
NK cells
38.9
PE CD56
BV785 CD16
CD8
17.4
CD4
80.9
SSC-H
BV570 CD4
CD95+
38.5
CD95-
61.4
BV421 CD95
BV570 CD4
TEM 31.1
TCM 48.8
TEMRA 5.10
TSCM 15.0
APC-Cy7 CD45RO
PE-D594 CCR7
Naive Cells
88.7
APC-Cy7 CD45RO
PE-D594 CCR7
CD95+
61.3
CD95-
38.7
BV421 CD95
BV570 CD4
TEM 27.5
TCM 6.07
TEMRA 51.3
TSCM 15.1
APC-Cy7 CD45RO
PE-D594 CCR7
Naive Cells
56.5
APC-Cy7 CD45RO
PE-D594 CCR7
NK cells
CD4
CD8
IFNg 2.12
PE-Cy7 (Y) IFNg
Alexa700 CD3
CD107a 3.28
PerCP-Cy5.5 CD107a
Alexa700 CD3
Q1 2.60
Q2 5.70
Q4 26.5
Q3 65.2
APC HLA-DR
BV650 CD38
Q5 1.45
Q6 0
Q8 98.5
Q7 0.024
BV605 Ki67
Ax488 Bcl2
IFNg 0.21
PE-Cy7 (Y) IFNg
Alexa700 CD3
CD107a 0.050
PerCP-Cy5.5 CD107a
Alexa700 CD3
IFNg 2.12
PE-Cy7 (Y) IFNg
Alexa700 CD3
CD107a 0.44
PerCP-Cy5.5 CD107a
Alexa700 CD3
CD4
CD8
Effector Mem
Q1 3.03
Q2 0
Q4 97.0
Q3 8.81E-3
BV605 Ki67
Ax488 Bcl2
Q5 19.4
Q6 2.69
Q8 74.7
Q7 3.19
APC HLA-DR
BV650 CD38
Q1 4.12
Q2 0
Q4 95.9
Q3 0.029
BV605 Ki67
Ax488 Bcl2
Q5 23.0
Q6 8.20
Q8 66.3
Q7 2.49
APC HLA-DR
BV650 CD38
Central Mem
Q1 0.84
Q2 0
Q4 99.2
Q3 2.80E-3
BV605 Ki67
Ax488 Bcl2
Q5 2.76
Q6 0.88
Q8 90.4
Q7 5.96
APC HLA-DR
BV650 CD38
Q1 1.72
Q2 0
Q4 98.3
Q3 0
BV605 Ki67
Ax488 Bcl2
Q5 7.00
Q6 3.50
Q8 86.4
Q7 3.10
APC HLA-DR
BV650 CD38
Stem Mem
Q1 0.41
Q2 0
Q4 99.6
Q3 9.12E-3
BV605 Ki67
Ax488 Bcl2
Q5 1.07
Q6 0.51
Q8 79.7
Q7 18.7
APC HLA-DR
BV650 CD38
Q1 1.32
Q2 0
Q4 98.7
Q3 0
BV605 Ki67
Ax488 Bcl2
Q5 1.59
Q6 1.56
Q8 88.1
Q7 8.78
APC HLA-DR
BV650 CD38
Temra
Q1 0.99
Q2 0
Q4 99.0
Q3 0
BV605 Ki67
Ax488 Bcl2
Q5 8.08
Q6 0.64
Q8 88.8
Q7 2.52
APC HLA-DR
BV650 CD38
Q1 1.88
Q2 0
Q4 98.1
Q3 7.82E-3
BV605 Ki67
Ax488 Bcl2
Q5 22.6
Q6 4.89
Q8 69.5
Q7 3.01
APC HLA-DR
BV650 CD38
Naive Cells
Q5 0.025
Q6 0
Q8 100.0
Q7 3.87E-3
BV605 Ki67
Ax488 Bcl2
Q1 0.54
Q2 0.15
Q4 83.8
Q3 15.5
APC HLA-DR
BV650 CD38
Q1 0.90
Q2 0.25
Q4 97.4
Q3 1.48
APC HLA-DR
BV650 CD38
Q5 0.079
Q6 0
Q8 99.9
Q7 0.056
BV605 Ki67
Ax488 Bcl2

**Supplementary Figure 1. Gating strategy.**

Panel A: Myeloid innate immune cells; Panel B: T cells and NK cells. Data were derived from the first participant enrolled in the study on Day 1 post-challenge.

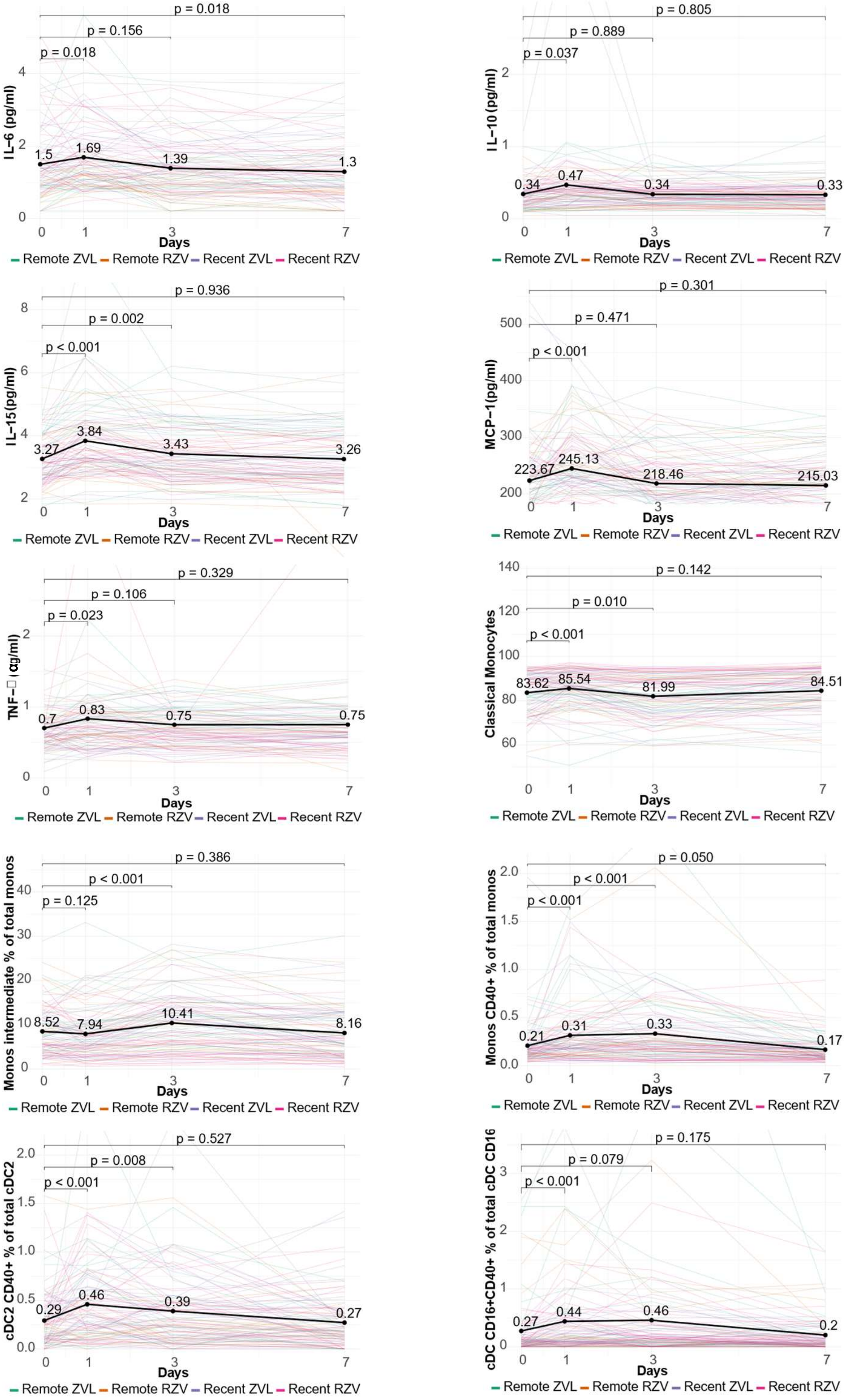

p = 0.018
p = 0.156
p = 0.018
IL−6 (pg/ml)
1.5
1.69
1.39
1.3
Days
Remote ZVL Remote RZV Recent ZVL Recent RZV
p = 0.805
p = 0.889
p = 0.037
IL−10 (pg/ml)
0.34
0.47
0.34
0.33
Days
Remote ZVL Remote RZV Recent ZVL Recent RZV
p = 0.936
p = 0.002
p < 0.001
IL−15(pg/ml)
3.27
3.84
3.43
3.26
Days
Remote ZVL Remote RZV Recent ZVL Recent RZV
p = 0.301
p = 0.471
p < 0.001
MCP−1(pg/ml)
223.67
245.13
218.46
215.03
Days
Remote ZVL Remote RZV Recent ZVL Recent RZV
p = 0.329
p = 0.106
p = 0.023
TNF− (αg/ml)
0.7
0.83
0.75
0.75
Days
Remote ZVL Remote RZV Recent ZVL Recent RZV
p = 0.142
p = 0.010
p < 0.001
Classical Monocytes
83.62
85.54
81.99
84.51
Days
Remote ZVL Remote RZV Recent ZVL Recent RZV
p = 0.386
p < 0.001
p = 0.125
Monos intermediate % of total monos
8.52
7.94
10.41
8.16
Days
Remote ZVL Remote RZV Recent ZVL Recent RZV
p = 0.050
p < 0.001
p < 0.001
Monos CD40+ % of total monos
0.21
0.31
0.33
0.17
Days
Remote ZVL Remote RZV Recent ZVL Recent RZV
p = 0.527
p = 0.008
p < 0.001
cDC2 CD40+ % of total cDC2
0.29
0.46
0.39
0.27
Days
Remote ZVL Remote RZV Recent ZVL Recent RZV
p = 0.175
p = 0.079
p < 0.001
cDC CD16+CD40+ % of total cDC CD16
0.27
0.44
0.46
0.2
Days
Remote ZVL Remote RZV Recent ZVL Recent RZV

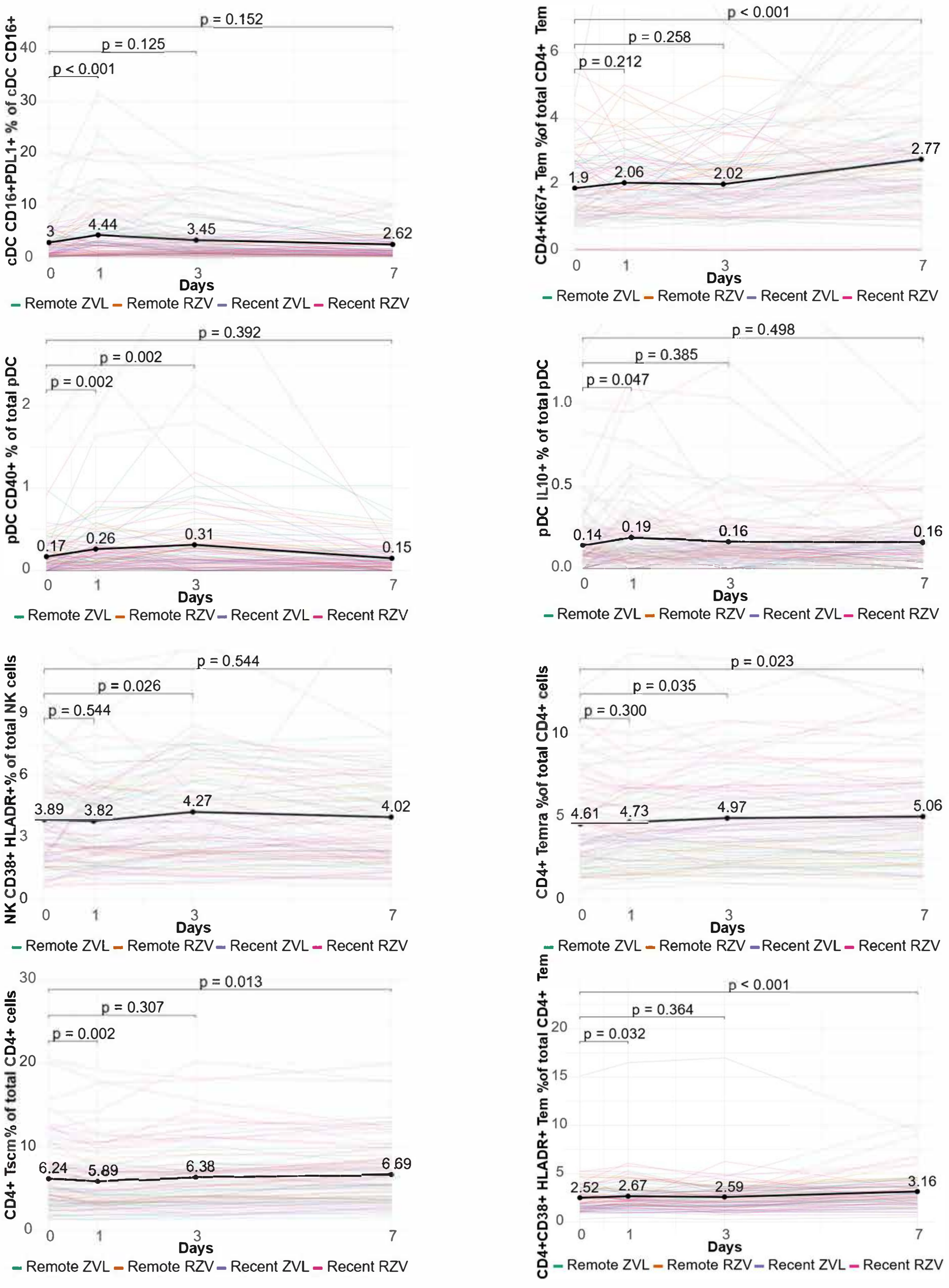


**Supplementary Figure 2. Pooled analysis of the systemic immune responses to the viral challenge.**

This figure complements Figure 3 by showing all the markers with significant frequency changes in circulating PBMC after the challenge that were not depicted in Figure 2. The data were derived from 88 participants across all vaccine groups with complete sets of data including all timepoints. Fine lines represent each participant and are individually color-coded for each vaccine group. The thick black line shows the average across all participants. The dots and numerical values on the thick line indicate means. The bars show comparisons among groups and the FDR-adjusted p values calculated by ANOVA for repeated measures.

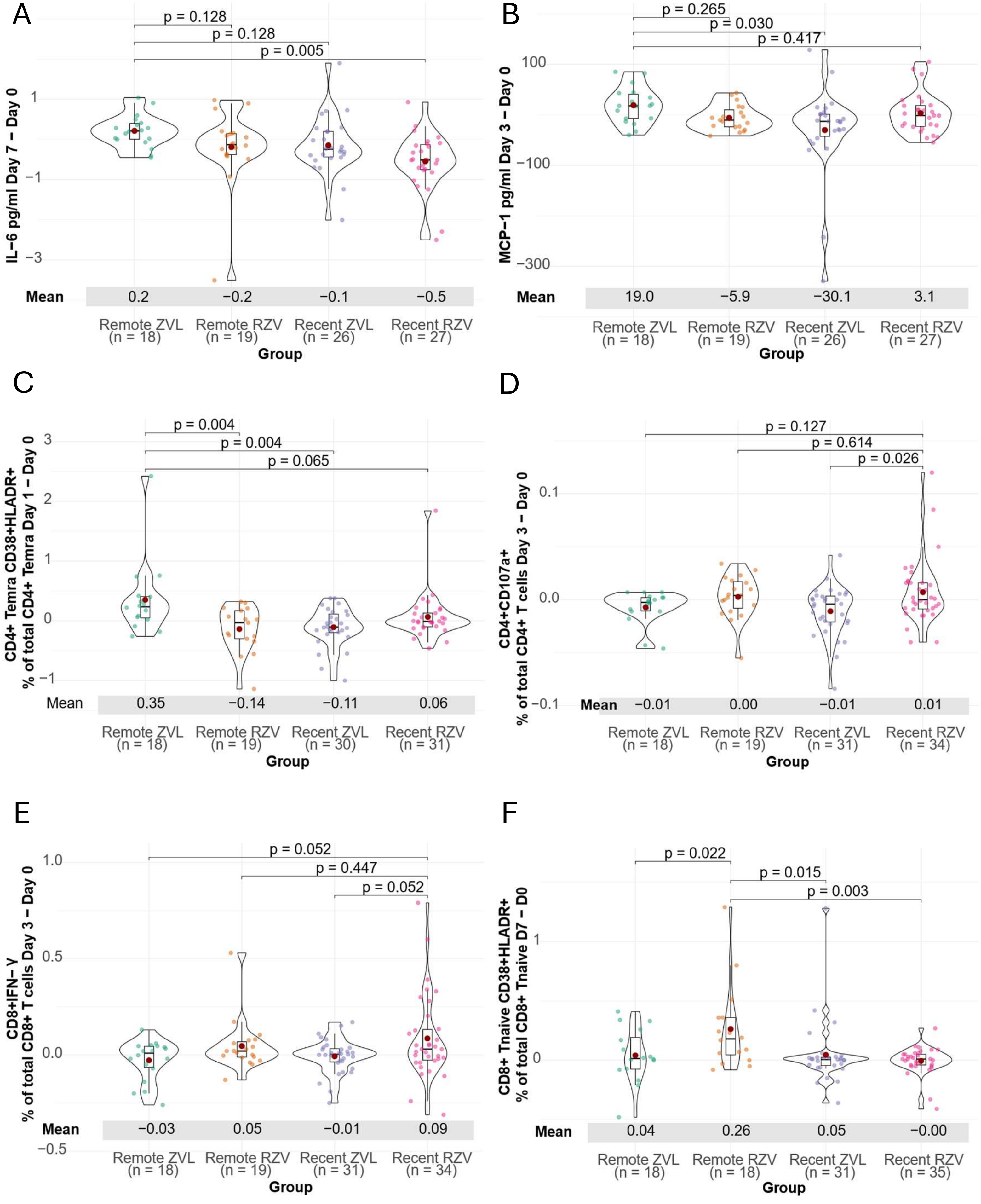
A
p = 0.128
p = 0.128
p = 0.005
IL−6 pg/ml Day 7 − Day 0
1
−1
−3
Mean
0.2
−0.2
−0.1
−0.5
Remote ZVL (n = 18)
Remote RZV (n = 19)
Recent ZVL (n = 26)
Recent RZV (n = 27)
Group
B
p = 0.265
p = 0.030
p = 0.417
MCP−1 pg/ml Day 3 − Day 0
100
−100
−300
Mean
19.0
−5.9
−30.1
3.1
Remote ZVL (n = 18)
Remote RZV (n = 19)
Recent ZVL (n = 26)
Recent RZV (n = 27)
Group
C
p = 0.004
p = 0.004
p = 0.065
CD4+ Temra CD38+HLADR+
% of total CD4+ Temra Day 1 − Day 0
3
2
1
0
−1
Mean
0.35
−0.14
−0.11
0.06
Remote ZVL (n = 18)
Remote RZV (n = 19)
Recent ZVL (n = 30)
Recent RZV (n = 31)
Group
D
p = 0.127
p = 0.614
p = 0.026
CD4+CD107a+
% of total CD4+ T cells Day 3 − Day 0
0.1
0.0
−0.1
Mean
−0.01
0.00
−0.01
0.01
Remote ZVL (n = 18)
Remote RZV (n = 19)
Recent ZVL (n = 31)
Recent RZV (n = 34)
Group
E
p = 0.052
p = 0.447
p = 0.052
CD8+IFN−Y
% of total CD8+ T cells Day 3 − Day 0
1.0
0.5
0.0
−0.5
Mean
−0.03
0.05
−0.01
0.09
Remote ZVL (n = 18)
Remote RZV (n = 19)
Recent ZVL (n = 31)
Recent RZV (n = 34)
Group
F
p = 0.022
p = 0.015
p = 0.003
CD8+ Tnaive CD38+HLADR+
% of total CD8+ Tnaive D7 − D0
1
0
Mean
0.04
0.26
0.05
−0.00
Remote ZVL (n = 18)
Remote RZV (n = 18)
Recent ZVL (n = 31)
Recent RZV (n = 35)
Group

**Supplementary Figure 3. Differential immune responses to VZV vOka replication among vaccine groups.**

The figure is complementary to Figure 4. It shows circulating immune parameters with differential increases post-challenge across vaccine groups. Increases in blood immune parameters from pre-challenge to Days 1, 3 and 7 post-challenge were compared among vaccine groups using data from 102 participants with paired samples that included Day 0. The dots represent individual differences from pre-challenge to the timepoint indicated on the y axis in each participant. Vaccine groups are color-coded. In the center of the violin plots, the boxes show medians, quartiles and ranges and the brown dots indicate means. Numerical values of the means are shown below the x axis. Above the horizontal bars between vaccine groups, FDR-adjusted p values calculated by ANOVA show the significance of the differences. Panels A, B, C show parameters with highest increases in the Remote ZVL group. Panels D, E show parameters with highest increases in the Recent RZV group. Panel F shows a parameter with highest increases in the Remote RZV group.

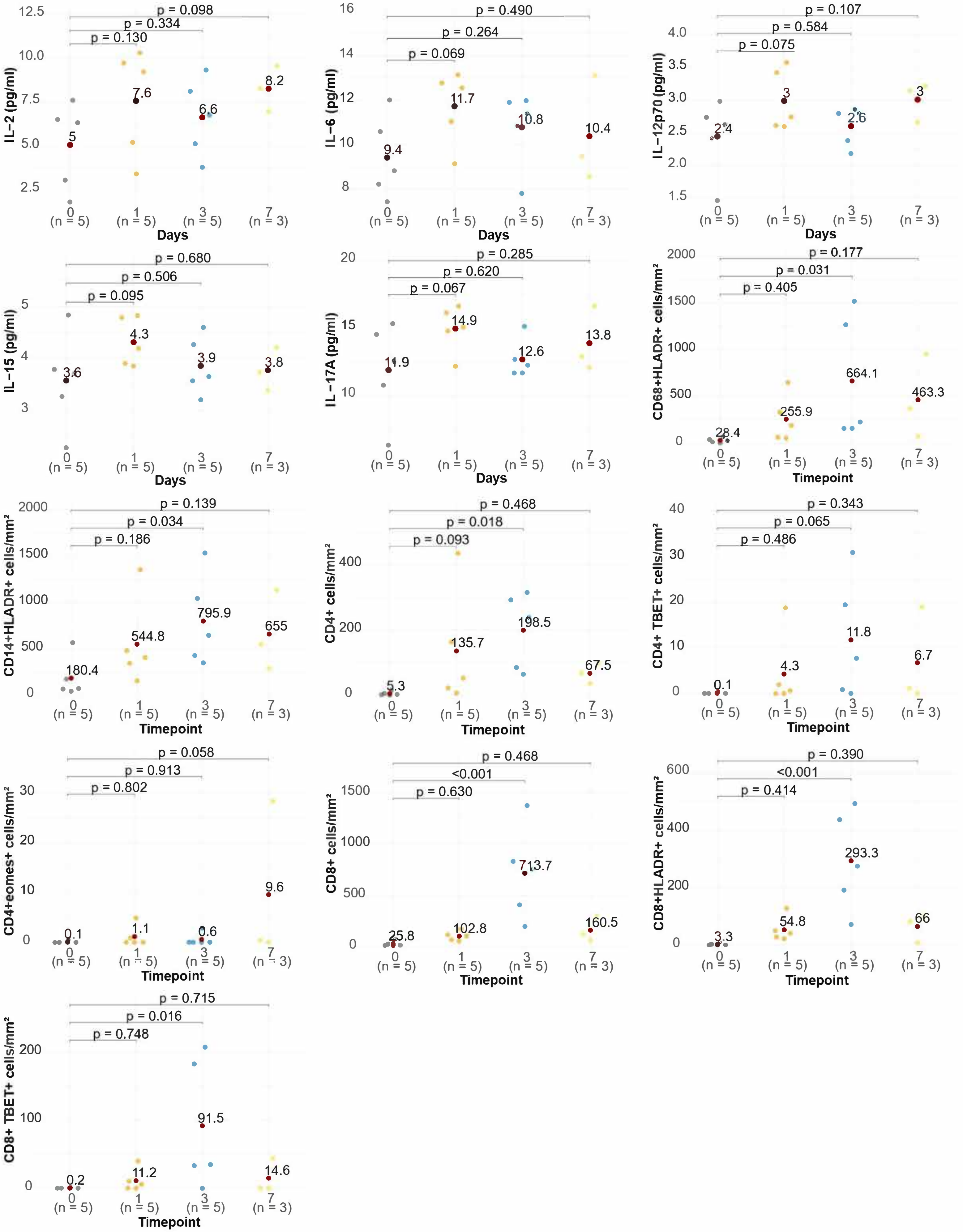

IL−2 (pg/ml)
p = 0.098
p = 0.334
p = 0.130
5
7.6
6.6
8.2
0 (n = 5)
1 (n = 5)
3 (n = 5)
7 (n = 3)
Days
IL−6 (pg/ml)
p = 0.490
p = 0.264
p = 0.069
9.4
11.7
10.8
10.4
IL−12p70 (pg/ml)
p = 0.107
p = 0.584
p = 0.075
2.4
3
2.6
3
IL−15 (pg/ml)
p = 0.680
p = 0.506
p = 0.095
3.6
4.3
3.9
3.8
IL−17A (pg/ml)
p = 0.285
p = 0.620
p = 0.067
11.9
14.9
12.6
13.8
CD68+HLADR+ cells/mm²
p = 0.177
p = 0.031
p = 0.405
28.4
255.9
664.1
463.3
Timepoint
CD14+HLADR+ cells/mm²
p = 0.139
p = 0.034
p = 0.186
180.4
544.8
795.9
655
CD4+ cells/mm²
p = 0.468
p = 0.018
p = 0.093
5.3
135.7
198.5
67.5
CD4+ TBET+ cells/mm²
p = 0.343
p = 0.065
p = 0.486
0.1
4.3
11.8
6.7
CD4+eomes+ cells/mm²
p = 0.058
p = 0.913
p = 0.802
0.1
1.1
0.6
9.6
CD8+ cells/mm²
p = 0.468
<0.001
p = 0.630
25.8
102.8
713.7
160.5
CD8+HLADR+ cells/mm²
p = 0.390
<0.001
p = 0.414
3.3
54.8
293.3
66
CD8+ TBET+ cells/mm²
p = 0.715
p = 0.016
p = 0.748
0.2
11.2
91.5
14.6

**Supplementary Figure 4. Immune responses at the cutaneous site of VZV vOka replication in Remote ZVL recipients.**

Data were derived from 18 participants. This figure is complementary to Figure 6. The graphs show cytokines, chemokines and cell subsets that significantly increased after challenge defined by $p<0.1$. Analyses were performed with ANOVA and p values show differences from pre-challenge to post-challenge timepoints.

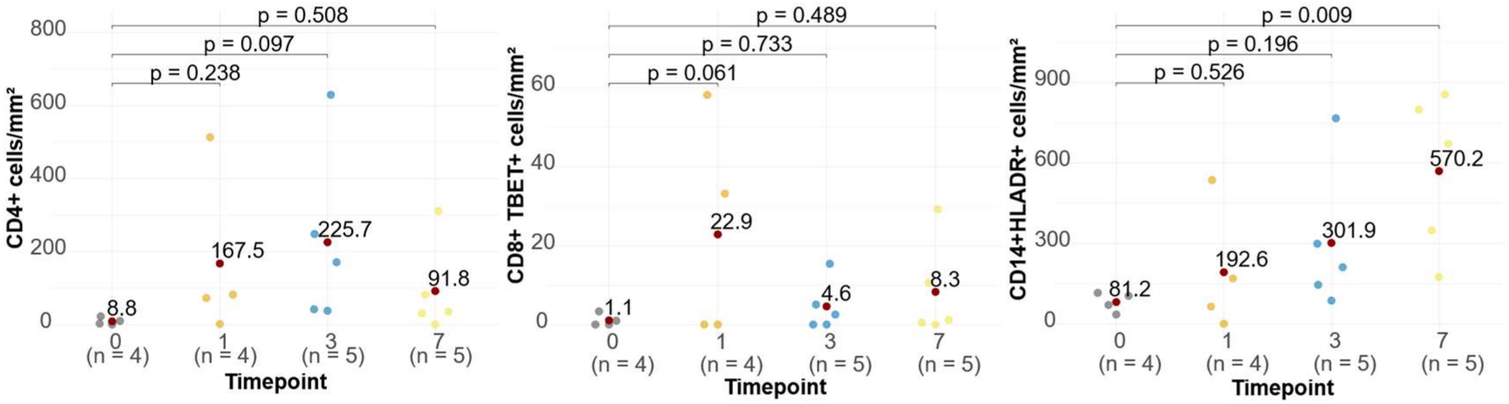


**Supplementary Figure 5. Immune responses at the cutaneous site of VZV vOka replication in Remote RZV recipients.**

Data were derived from 18 participants. This figure is complementary to Figure 6. The graphs show cell subsets that significantly increased after challenge defined by ANOVA FDR-adjusted p<0.1. p values on the graph were calculated post-hoc and show differences from pre-challenge to post-challenge timepoints.

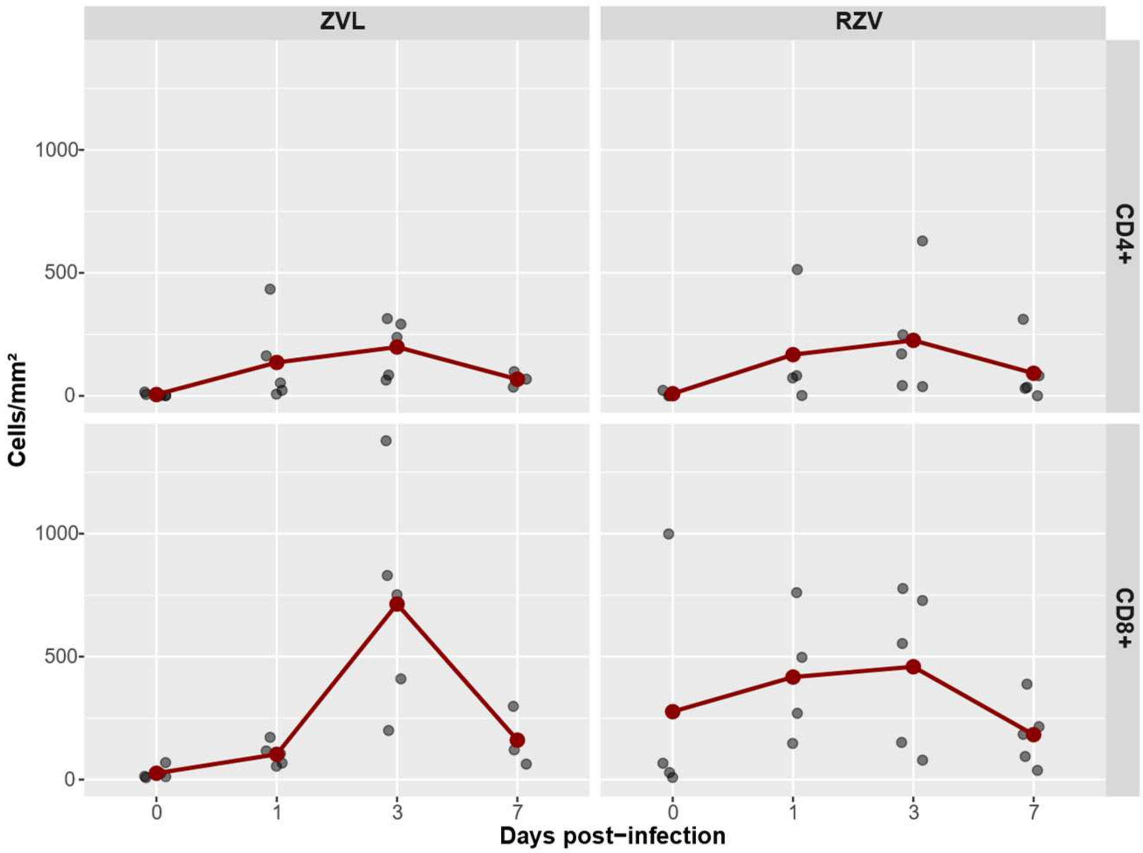


**Supplementary Figure 6. Kinetics of CD4+ and CD8+ T cells in the skin.**

This figure is complementary to Figure 7. The graph shows CD4+ and CD8+ T cell densities before infection and at the site of infection at each timepoint post-challenge. The grey dots show individual participant data. The brown dots represent means. The line connecting the means emphasizes the differential kinetics in Remote RZV and Remote ZVL recipients.
